# Modeling the impact of school reopening and contact tracing strategies on COVID-19 dynamics in different epidemiologic settings in Brazil

**DOI:** 10.1101/2021.10.22.21264706

**Authors:** Marcelo Eduardo Borges, Leonardo Souto Ferreira, Silas Poloni, Angela Maria Bagattini, Caroline Franco, Michelle Quarti Machado da Rosa, Lorena Mendes Simon, Suzi Alves Camey, Ricardo de Souza Kuchenbecker, Paulo Inácio Prado, José Alexandre Felizola Diniz Filho, Roberto André Kraenkel, Renato Mendes Coutinho, Cristiana Maria Toscano

## Abstract

Among the various non-pharmaceutical interventions implemented in response to the COVID-19 pandemic during 2020, school closures have been in place in several countries to reduce infection transmission. Nonetheless, the significant short and long-term impacts of prolonged suspension of in-person classes is a major concern. There is still considerable debate around the best timing for school closure and reopening, its impact on the dynamics of disease transmission, and its effectiveness when considered in association with other mitigation measures. Despite the erratic implementation of mitigation measures in Brazil, school closures were among the first measures taken early in the pandemic in most of the 27 states in the country. Further, Brazil delayed the reopening of schools and stands among the countries in which schools remained closed for the most prolonged period in 2020. To assess the impact of school reopening and the effect of contact tracing strategies in rates of COVID-19 cases and deaths, we model the epidemiological dynamics of disease transmission in 3 large urban centers in Brazil under different epidemiological contexts. We implement an extended SEIR model stratified by age and considering contact networks in different settings – school, home, work, and elsewhere, in which the infection transmission rate is affected by various intervention measures. After fitting epidemiological and demographic data, we simulate scenarios with increasing school transmission due to school reopening.

Our model shows that reopening schools results in a non-linear increase of reported COVID-19 cases and deaths, which is highly dependent on infection and disease incidence at the time of reopening. While low rates of within-school transmission resulted in small effects on disease incidence (cases/100,000 pop), intermediate or high rates can severely impact disease trends resulting in escalating rates of new cases even if other interventions remain unchanged. When contact tracing and quarantining are restricted to school and home settings, a large number of daily tests is required to produce significant effects of reducing the total number of hospitalizations and deaths. Our results suggest that policymakers should carefully consider the epidemiological context and timing regarding the implementation of school closure and return of in-person school activities. Also, although contact tracing strategies are essential to prevent new infections and outbreaks within school environments, our data suggest that they are alone not sufficient to avoid significant impacts on community transmission in the context of school reopening in settings with high and sustained transmission rates.

## 1. Introduction

Among the various non-pharmaceutical interventions (NPIs) recommended to mitigate the current COVID-19 pandemic, school closure is the most commonly implemented and for the longest period among low and middle-income countries (LMIC). The additional education and development burden resulting from prolonged school closure is of particular concern, especially when this strategy has been prioritized over other mitigation measures that have been consistently demonstrated and recommended as first-line actions (Panovska-Griffiths et al., 2020). As such, considering its far-reaching consequences, it has been recommended that school closures are the last NPI measure to be implemented and the first one to be lifted (Van Lancker and Parolin, 2020; World Health Organization et al., 2020).

The Brazilian population is experiencing an unprecedented health crisis due to the COVID-19 pandemic. Previously existing health structure and health system inequities in the country have been aggravated by the pandemic. In addition, epidemiologic, socio-economic, geographical, and political challenges including uncoordinated actions, variability of public health response among the various states, delayed and insufficient implementation of NPIs, among others (Silva et al., 2020), coupled with a limited preparedness and emergency response system in place (Idrovo et al., 2021), have further impacted the ability to mitigate the pandemic, resulting in one of the highest COVID-19 burdens in the world. With an estimated population of 211 million and being the fifth largest country in territorial extension, Brazil presented, as of April 2021, the second-largest number of COVID-19 deaths worldwide (Pan American Health Organization, 2020). As a result of the COVID-19 pandemic, a decline of 1.94 years in life expectancy at birth has been estimated (Castro et al., 2021).

Today, more than a year after the onset of the pandemic, Brazil ranks first in the world on the duration of school closure (UNICEF, 2021), having implemented strict school closure policies early on in the pandemic and having delayed its reopening (Reimers and Schleicher, 2020).

Dynamic transmission modelling has provided evidence to support decision-making related to the timing and impact of various NPI measures, among others (Aguas et al., 2020; Anirudh, 2020; Davies et al., 2020; Panovska-Griffiths et al., 2021). It is recommended that school reopening is followed by large-scale, population-wide testing of symptomatic individuals and effective contact tracing of related contacts, followed by isolation of diagnosed individuals and quarantine of contacts. In Brazil, these large-scale diagnostic testing and contact tracing strategies have remained limited throughout the pandemic (Panovska-Griffiths et al., 2020).

Considering its continental dimensions and regional specificities, and the scenario outlined above, modelling the dynamics of SARS-CoV-2 transmission in selected states in Brazil’s considering school reopening following the first wave of the epidemic provides an opportunity to evaluate and provide additional evidence to support policymaking in this regard. Hence, we have modelled the impact of school reopening during the COVID-19 pandemic, aiming to identify optimal timing, epidemiological indicators, and additional strategies such as contact tracing, which may allow for safer reopening in three major urban centers in Brazil mimicking LMIC settings. These results shed light on future evidence-based policy implementation to mitigate the ongoing COVID-19 pandemic in LMIC.

### OBJECTIVES

Through dynamic transmission modelling, the study objectives are: a) to project the impact of schools’ reopening in COVID-19 cases and deaths, b) to evaluate the impact of contact tracing strategies during school reopening and whether these could be used to mitigate transmission in the school environment and enable a safer school reopening, c) to identify epidemiological indicators related to the timing of school reopening that are associated with additional disease burden, and d) to contrast the projected impact of school reopening and the true incidence of COVID-19 cases and deaths in 2021 in Brazil.

## 2. Material and Methods

In the following subsections, we first present information on each of the study sites considered in modelling, including demographic and COVID-19 epidemiologic indicators. Next, we provide details of the dynamic transmission model used, the contact tracing and case isolation strategies considered, present model parameters and sources considered, and describe the NPIs considered in the model. Finally, we describe the data collection and model calibration processes, explain the school reopening scenarios considered, and describe sensitivity analyses conducted. We followed the recommendations of the International Society for Pharmacoeconomics and Outcomes Research (ISPOR) and the Society of Medical Decision Making (SMDM) Modeling Good Research Practices Task Force for dynamic transmission modeling (Pitman et al., 2012).

### 2.1 Study sites

Brazil is an upper-middle-income country with more than 200 million inhabitants, occupying a vast area divided into 5 macro-regions, 27 states, and 5,570 municipalities with significant variation of socio-economic, demographic, and geographic patterns. The model considered 3 municipalities which are state capitals, each of them located in different macro-regions of the country - the city of São Paulo-SP in the southeastern region, the city of Porto Alegre-RS in the south region, and the city of Goiânia-GO in the central-west region. São Paulo is the most populous city in South America, has the largest GDP in the country, and is where the first case of COVID-19 was notified in Brazil on February 26^th^, 2020. Goiânia-GO and Porto Alegre-RS are medium-sized capitals (**Table 1**).

**Table 1.**
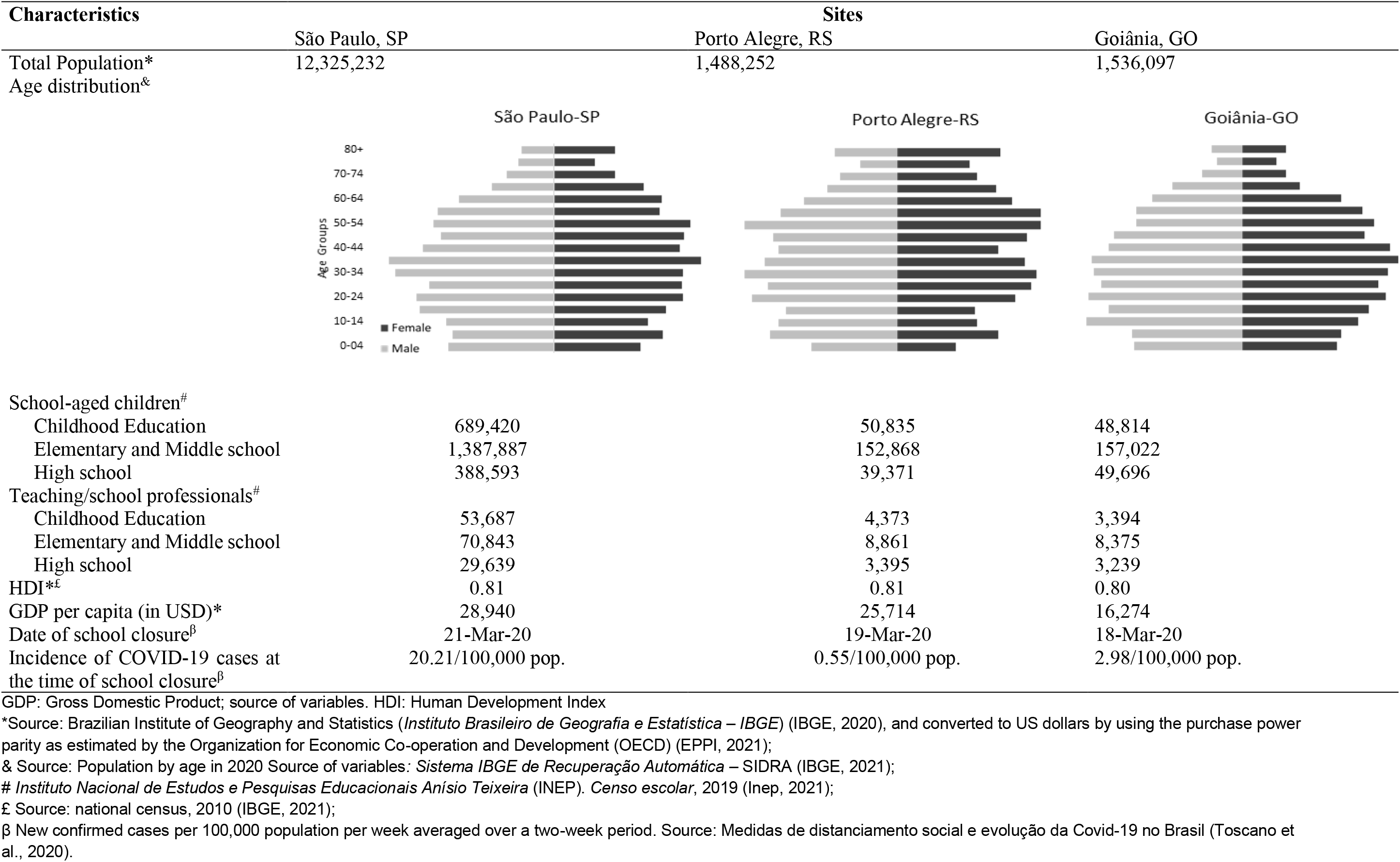
Epidemiological and socio-demographic characterization of study sites. Brazil, 2020.

| Characteristics | Sites |  |  |
| --- | --- | --- | --- |
|  | São Paulo, SP | Porto Alegre, RS | Goiânia, GO |
| Total Population* | 12,325,232 | 1,488,252 | 1,536,097 |
| Age distribution<sup>&</sup> |  |  |  |
| School-aged children<sup>#</sup> |  |  |  |
| Childhood Education | 689,420 | 50,835 | 48,814 |
| Elementary and Middle school | 1,387,887 | 152,868 | 157,022 |
| High school | 388,593 | 39,371 | 49,696 |
| Teaching/school professionals<sup>#</sup> |  |  |  |
| Childhood Education | 53,687 | 4,373 | 3,394 |
| Elementary and Middle school | 70,843 | 8,861 | 8,375 |
| High school | 29,639 | 3,395 | 3,239 |
| HDI*<sup>£</sup> | 0.81 | 0.81 | 0.80 |
| GDP per capita (in USD)* | 28,940 | 25,714 | 16,274 |
| Date of school closure<sup>β</sup> | 21-Mar-20 | 19-Mar-20 | 18-Mar-20 |
| Incidence of COVID-19 cases at the time of school closure<sup>β</sup> | 20.21/100,000 pop. | 0.55/100,000 pop. | 2.98/100,000 pop. |

GDP: Gross Domestic Product; source of variables. HDI: Human Development Index
\*Source: Brazilian Institute of Geography and Statistics (*Instituto Brasileiro de Geografia e Estatística – IBGE*) (IBGE, 2020), and converted to US dollars by using the purchase power parity as estimated by the Organization for Economic Co-operation and Development (OECD) (EPPI, 2021);
& Source: Population by age in 2020 Source of variables: *Sistema IBGE de Recuperação Automática – SIDRA* (IBGE, 2021);

With the onset of the COVID-19 pandemic in 2020, public and private schools were closed in mid-late March, with in-person school activities suspended and shifting to remote online activities, including childhood, elementary, high school, and college education settings. School reopening has been postponed on several occasions, varying over time and by state. The city of Porto Alegre authorized partial school return at the end of September 2020, with little adherence from schools and students. In São Paulo, school reopening was authorized in October 2020]. In Goiânia, the reopening was initially authorized for private schools and early childhood education only, and later in November expanded to public schools and all levels of education. In all locations, strict sanitary protocols were in place and a reduced number of students were allowed to be present, requiring rotations among students. The dates schools were closed and reopened, as well as data on COVID-19 cases and deaths over time, by State, are publicly available at https://medidas-covidbr-iptsp.shinyapps.io/painel/.

### 2.2 Epidemiological Model

We have developed an extended age-structured SEIR compartmental model based on (Franco et al., 2021) that accounts for different levels of disease severity (Figure 1), also accounting for case isolation and quarantine of contacts (Figures 1 and 2). The force of SARS-Cov-2 infection transmission is affected by the estimated reduction of contact for each NPI and adherence of NPIs in place, also considered in the model. Model compartments are represented in Figure 1 and a detailed description of the model on the supplementary material, section 2.

**Figure 1.**
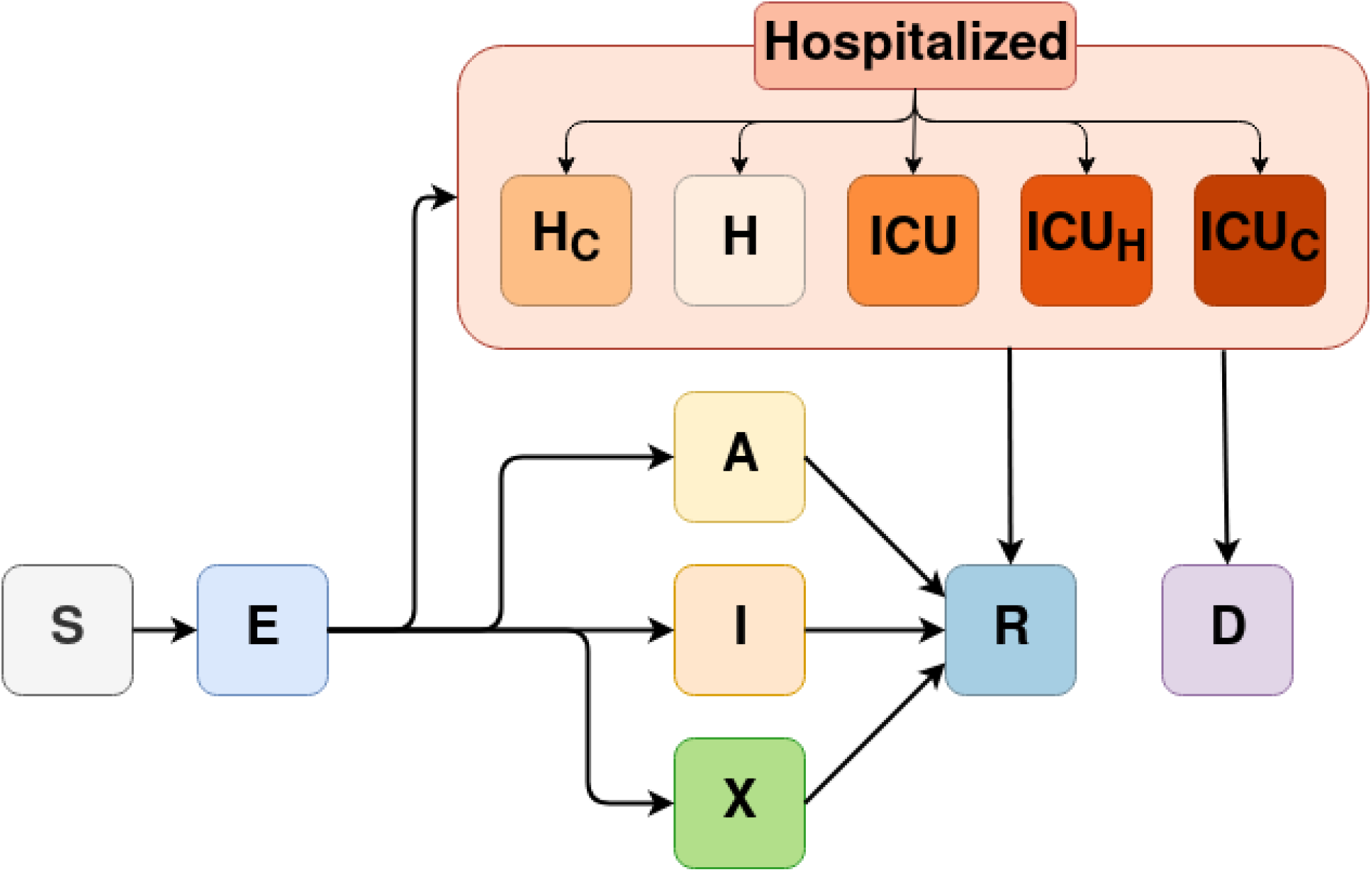
Diagram of the mathematical model and its compartments. Solid arrows describe the possible pathways of individuals on the susceptible (S) compartment after exposure to infection (E), including asymptomatic infection (A), symptomatic infection (I), and cases requiring hospitalization in regular hospital wards (H) or intensive care unit beds (ICU). If the requirement for hospitalization exceeds the health system capacity, individuals with severe disease move to the compartments of unattended cases requiring hospitalization (H_c_), unattended cases requiring ICU (ICU_h_), or cases requiring ICU treated on hospital wards (ICU_c_). All infected individuals can either recover (R) or die (D).

**Figure 2.**
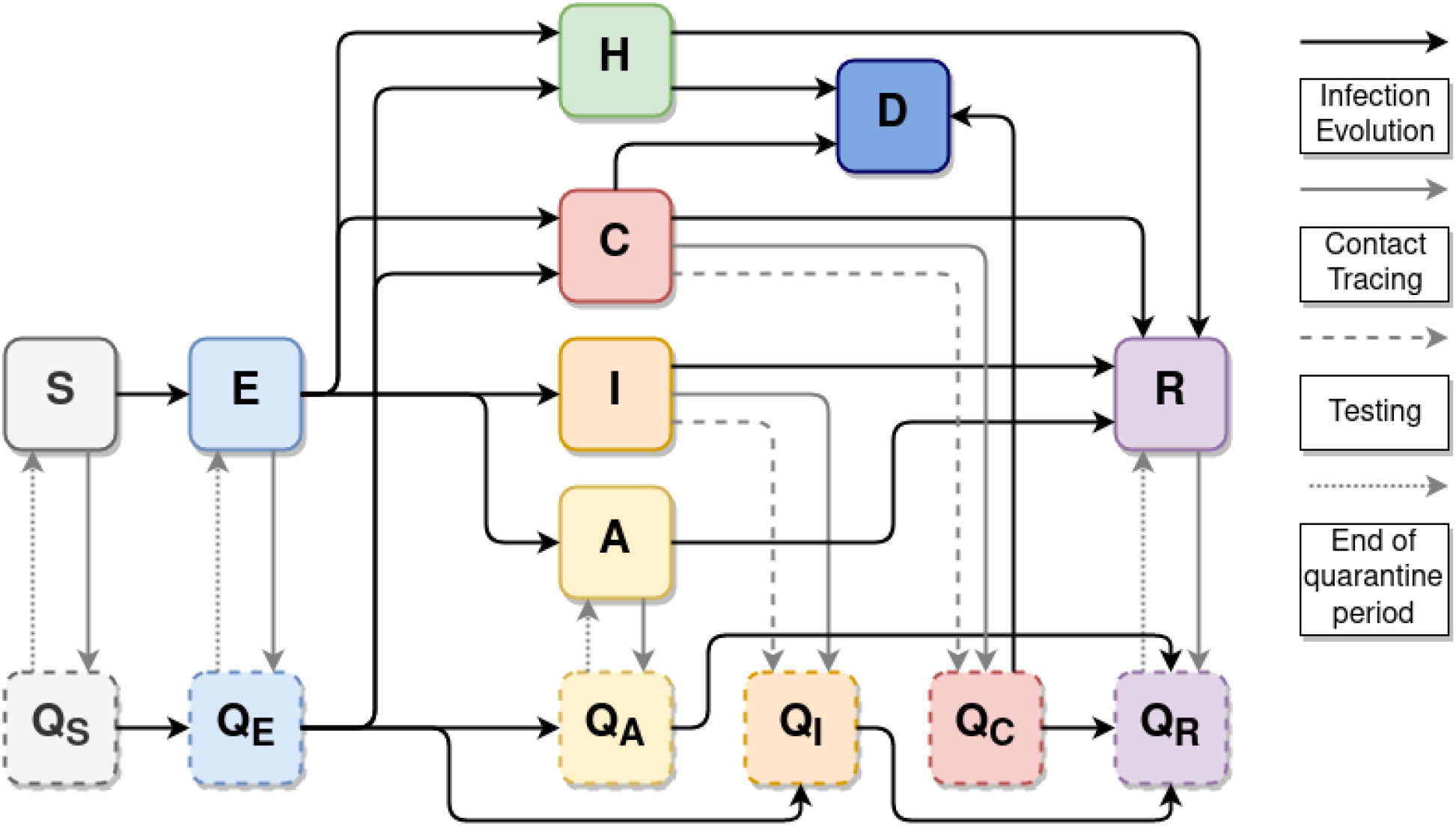
Contact tracing flow diagram. H comprises all hospitalized compartments (*H, ICU, ICU*_*h*_), and C comprises all critical compartments (*H*_*c*_, *ICU*_*c*_) who have not received attendance. Solid black arrows describe the infection pathways in the model, as in Figure 1. Solid gray arrows describe individuals who are quarantined through contact tracing. Dashed gray arrows describe individuals who are isolated/quarantined because of positive testing in an asymptomatic individual. Dotted gray arrows describe the pathway of individuals after the quarantine period.

From the epidemiological perspective, the progression of infection status and transmission among individuals occurs within different compartments as follows: the susceptible (S) compartment includes individuals without previous exposure to SARS-Cov-2 infection. Once becoming infected, exposed individuals transition to the pre-symptomatic (E) compartment, presenting a relative level of infectiousness. Once fully infectious (I), individuals may be still asymptomatic (A) or become symptomatic (I). Regardless of being symptomatic or asymptomatic, a fraction of individuals may self-isolate themselves (X) to decrease the risk of infecting others. Individuals from the asymptomatic (A), symptomatic (I), and self-isolated (X) compartments will transition to the Recovered (R) compartment after the end of the infectious period. Alternatively, infected individuals with more severe disease (ie., requiring hospitalization) transition to the hospitalization compartments, which include either hospitalization in regular hospital wards (H) or intensive care units (ICU). If, as in the case of a health system collapse, the demand for hospital and ICU beds exceeds the number of available beds, unattended individuals transition to the compartments H_c_ (unattended cases requiring hospitalization) and ICU_c_ (unattended cases requiring ICU), respectively. Furthermore, those requiring ICU beds can be managed in hospital beds (ICU_h_) if that is the only option available. Individuals in those compartments will transition to the Recovered (R) or Deceased (D) compartments. We assume that all deaths due to COVID-19 will occur in hospitalized individuals and that no deaths occur in symptomatic individuals not requiring hospitalization.

We considered population strata divided into 5 year age sub-groups, ranging from 0 to 95 years and over. Contacts among individuals can occur in four different settings: work, home, school, and in the community (i.e. public transportation, social gatherings, shopping activities, etc.). The contact rate between individuals by age groups and by settings type are based on the contact matrices (Prem et al., 2021), considering the Brazilian population in urban environments (see supplementary material, section 2). The infection transmission probability is derived from the pattern of social contacts, and the frequency of potential contacts, varying by age group and the setting where the contact occurs.

In addition, the infection transmission rate can be affected by the following NPIs considered in the model: self-isolation, social distancing, use of masks, home-office, cocooning of older adults, and school closure (see supplementary material, section 3).

### 2.3 Model parameters, data source, and fitting

Model parameters considered national information systems and the best available evidence from the literature (Table 2, see also the supplementary material, section 4). Upon the lack of data from the literature, some of the parameters were assumed based on reasonable values.

**Table 2.** Model parameters considered in the analysis of COVID-19 school reopening in Brazil, 2020.

| **Symbol** | **Description** | **Value** | **Source** |
| --- | --- | --- | --- |
| **rho** | Relative infectiousness of incubation phase | 10.5% | (W. E. Wei et al., 2020) |
| **rhos** | Relative percentage of regular daily contacts when hospitalized | 10% | Assumed |
| **omega** | Average duration of immunity | ∞ | Assumed |
| **gamma<sup>-1</sup>** | Average of incubation period | 5.8 days | (Y. Wei et al., 2020) |
| **nui<sup>-1</sup>** | Average duration of the symptomatic infection period | 9 days | (Cevik et al., 2021) |
| **pclin** | Probability upon infection of developing clinical symptoms by age groups |  |  |
|  | 0-19 years | 30.5% | (São Paulo, 2020) |
|  | 20-59 years | 56% | (Sun et al., 2020) |
|  | 60 or more years | 69% | (Sun et al., 2020) |
| **scale_ihr** | Scaling factor for infection hospitalization rate | 0.8 | Fitting |
| **mask eff** | Estimated reduction of contact due to mask use | 85% | (Chu et al., 2020) |
| **selfis eff** | Estimated reduction of contact due to self-isolation if symptomatic | 80% | Assumed |
| **mask cov** | Adherence to mask usage | Varies by study site and over time | See SM sections 3, 4, and 5 |
| **selfis cov** | Adherence to self-isolation | Varies by study site and over time | See SM sections 3, 4, and 5 |
| **dist cov** | Adherence to social distancing at the community level | Varies study site and over time | See SM sections 3, 4, and 5 |
| **work cov** | Adherence to home-office | Varies study site and over time | See SM sections 3, 4, and 5 |
| **School cov** | Adherence to online (not in-person) school activities | Varies study site and over time | See SM sections 3, 4, and 5 |
| **cocoon_cov** | Adherence to cocooning of older adults | Varies study site and over time | See SM sections 3, 4, and 5 |
| **dist eff** | Reduction of contacts in the community among those adhering to social distancing | 95% | Assumed |
| **work eff** | Reduction of contacts at work among those adhering to home-office | 95% | Assumed |
| **school eff** | Reduction of contacts in school upon school closure | 100% | Assumed |
| **cocoon eff** | Reduction of contacts with older adults in all settings as a result of cocooning older adults | 90% | Assumed |
| **report** | Percentage of all asymptomatic infections that are reported | 0% | SIVEP (DATASUS, 2020) |
| **reporth** | Percentage of non-severe hospitalizations that are appropriately treated | 95 | SIVEP (DATASUS, 2020) |
| **reportc** | Percentage of all symptomatic infections that are reported | 1% | SIVEP (DATASUS, 2020) |
| **give** | System capacity stressor | 65% | Assumed |

SM: Supplementary Material

The effectiveness of each NPIs considered in the model is dependent on the adherence (*NPI*_*cov (t)*_ **∈** {0,1}) and estimated reduction of contacts (*NPI*_*eff*_ **∈** {0,1}) for each intervention, each varying from 0 to 1 (0-100%). Adherence to each intervention by study site was based on the adherence of the population to each NPI in 2020 and was obtained from monitoring NPI implementation and level of strictness over time (Silva et al., 2020) (see supplementary material, sections 3-5). The reduction of contacts (in %) refers to the percentage of potential contacts in a given location that is prevented as a result of the intervention and was assumed as fixed over time (Table 2).

For each study site, length of stay in hospital wards and ICU, and Intra-Hospital Fatality Rate (IHFR) was obtained from Brazil’s Epidemiological Surveillance Information System for Acute Respiratory Illness (SIVEP) (DATASUS, 2020; Ranzani et al., 2021) (see further details on the supplementary material, section 4).

Fitting was based on the number of COVID-19 hospitalizations and deaths reported to the SIVEP database (see additional details on the supplementary material, section 4).

### 2.4 Time horizon and analytic framework

School reopening intervention in the model was set at February 1st, 2021, which was the scheduled date for reopening school in most Brazilian states, following summer holidays in December 2020 and January 2021. Incidence of COVID-19 cases and deaths was projected throughout 2021, until December.

### 2.5. School reopening scenarios

We evaluated the effect of reopening schools by simulating scenarios considering increasing values of the percentage of potential contacts of individuals in the school setting (henceforth described as PCS) after reopening schools. The scenarios consider the parameter “adherence to online (not in-person) school activities”, and results from (100 – **school cov** parameter), presented in percentage. This approach can reflect a combination of strategies that reduce the likelihood of contact and transmission in a school environment, including but not limited to implementation of hybrid learning approaches (online and in-person), and infection prevention measures when returning to school such as limiting the maximum number of students in the classroom, natural ventilation of indoor spaces, a distance of at least 6 feet between students and teachers/staff, mask use at school, among others. We implemented these scenarios by setting values ranging from 0 to 100% at regular intervals of 20, starting on February 1st, 2021.

To assess solely the effect of school reopening, we assumed that other interventions remained unchanged and compared the weekly incidence of new COVID-19 cases and deaths throughout the year 2021. We also evaluated the additional incidence of cases and deaths due exclusively to school reopening in three age groups: young (less than 20 years old) adults (between 20 and 59 years old), and older adults (more than 60 years old). This value was estimated by the difference in the cumulative incidence of cases and deaths on December 31, 2021, compared to the baseline scenario where schools remain closed for the whole year (PCS = 0).

### 2.6 Diagnostic testing, contact tracing, isolation, and quarantine scenarios

Considering WHO recommendations (World Health Organization, 2020), we also evaluated the effects of case isolation, contact tracing, and quarantining contacts in schools upon reopening (henceforth described as CT model), to mitigate transmission events within the school environment. This consists of identifying symptomatic individuals and their contacts, isolating positive cases, and quarantining contacts who tested positive for COVID-19 after diagnostic testing. Since in Brazil diagnostic testing is prioritized for symptomatic hospitalized individuals (Kameda et al., 2021), our contact tracing model prioritizes the usage of the available tests to severe and symptomatic individuals and, if any available, it proceeds to test asymptomatic individuals. In the model, contact tracing considers the various compartments of the model. The CT model is further described in the supplementary material, section 2.

Figure 2 presents the flow diagram of the model compartments and quarantined compartments considered. Symptomatic or asymptomatic individuals that had high-risk contact with the index cases are transferred to a quarantine compartment (Q, with the respective sub-index). In this compartment, contacts are restricted to their households. Symptomatic individuals are isolated until recovery, here depicted as quarantine of infected. Asymptomatic infected individuals were kept in quarantine until the end of the isolation/quarantine period (10 days). Note that regardless of the contact tracing strategy, symptomatic individuals may already self-isolate themselves according to the adherence to the self-isolation intervention.

As our main objective was to assess whether contact tracing strategies can contribute to mitigating infection transmission in the school environment, and potentially increase the safety of reopening schools, we only considered CT strategies restricted to the household and school environments.

Finally, for each scenario, we estimated the reduction in the final cumulative incidence of COVID-19 cases and deaths after the implementation of contact tracing and isolation strategies for the period of simulation. For each scenario of PCS, we simulated daily tests for the population proportional to each city’s population size, until increasing the number of tests no longer reduced the incidence of cases and deaths. The use of the contact tracing strategy starts on the same date where the school reopening starts in our model. We assumed that all individuals that became contacts are timely traced and isolated, thus, the effectiveness of the strategies depends on the number of daily tests available for use in the population.

### 2.7 Sensitivity analysis

To account for both parameter and model uncertainties, we performed a sensitivity analysis varying values for the following nine parameters: relative infectiousness of incubation phase (**rho**), relative percentage of regular daily contacts when hospitalized (**rhos**), probability upon infection of developing clinical symptoms for young individuals (**pclin young**), estimated reduction of contact due to mask use (**mask eff**), estimated reduction of contact due to self-isolation if symptomatic (**self eff**), adherence to self-isolation (**selfis cov**), adherence to social distancing at the community level (**dist cov**), adherence to home-office (**work cov**), and adherence to cocooning of older adults (**coccoon cov**). Since changes in the value of a given parameter evaluated by the sensitivity analysis require a new estimation for the free parameters of the model, we performed a new fitting including the parameter evaluated on the sensitivity analysis (see details on the supplementary material, section 5), for the same period used for the previous fitting in each city. We then used the new fitted values to simulate the school reopening scenarios and compared the additional incidence of cases and deaths due to school reopening relative to the model with the original values of the parameter chosen for the sensitivity analysis.

### 2.8 Contrasting modelled and true incidence of events after school reopening

As we modelled only school reopening with additional combined contact tracing strategies, without accounting for other epidemiologic patterns of disease progression which at that time were not amenable to modelling, particularly the emergence and circulation of variants of concern with different transmissibility patterns, and unanticipated relaxation of other NPIs by policy makers, we also contrasted the projected COVID-19 incident cases and deaths over time after school reopening, and the true epidemic curves of COVID-19 cases, hospitalization and deaths in the study sites, considering the same analytical horizon.

## 3. Results

### 3.1 School reopening

Our model predicts that school reopening results in an increase in the number of new COVID-19 cases and related deaths proportional to the degree of increase of potential contacts in the school environment (Figure 3). The estimated increase is affected by local epidemiological indicators, particularly the incidence and trend of COVID-19 cases and deaths, at the moment of reopening in each study site. As shown in Figure 3, in settings of high disease occurrence, the impact of school reopening is projected to be more significant.

**Figure 3.**
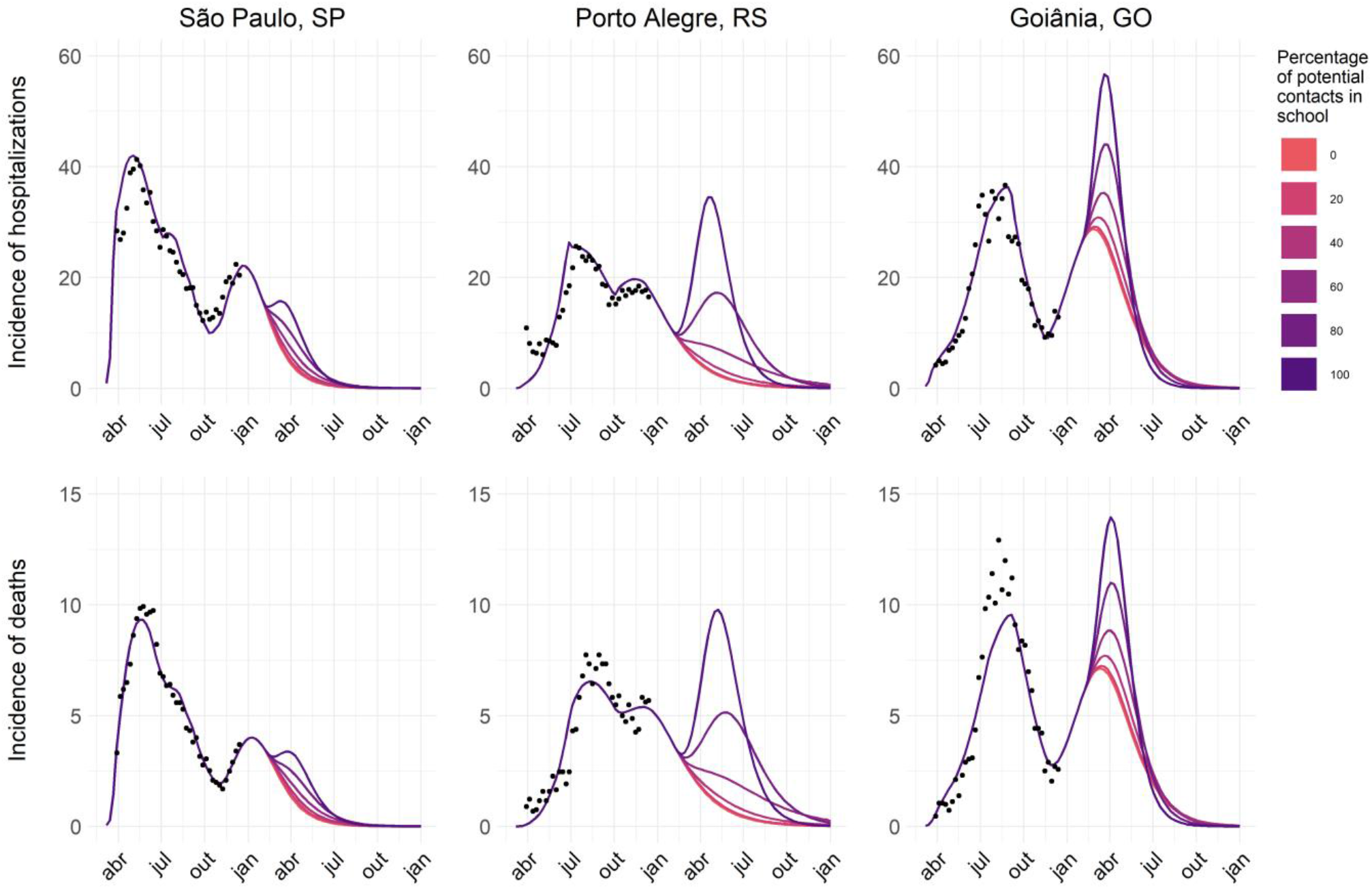
Epidemiological dynamics for different scenarios of increase in the percentage of potential contacts in school for three Brazilian capitals: São Paulo, Porto Alegre, and Goiânia. Colors represent the percentage of potential contacts of individuals in the school setting after reopening schools on February 1st, and dots represent the hospitalization and death data from the SIVEP database.

In a scenario where schools remain closed throughout 2021 and other interventions unchanged, both São Paulo and Porto Alegre were expected to present a decline in the number of cases with the epidemic under control after the second semester of that year. However, the rate of decrease in the number of cases is reduced on scenarios with a gradual increase in transmissibility in schools (for instance, PCS ⩽ 60% in Porto Alegre, and PCS ⩽ 80% in São Paulo). In scenarios where PCS exceeds these values, the effects of increasing transmission within schools can reverse the trend and lead to a third surge in the number of cases in both capitals. Goiânia, on the other hand, by the end of 2020 was experiencing a second surge in the number of cases. An increase in transmission in the school environment is predicted to sustain the growth of cases for a longer time proportional to PCS: while small PCS values (⩽ 40%) results in small increases in the number of cases, higher PCS values (> 60%) may lead to a daily number of cases and deaths close or even higher than observed in the first peak of cases.

School transmission also resulted in a non-linear increase in the excess cumulative incidence of COVID-19 cases and deaths in all study sites, affecting age groups disproportionately (Figure 4). The excess cumulative incidence represents the additional incident cases and deaths projected by the end of 2021, compared to the baseline scenario where schools remain closed throughout the year, computed for each age group.

**Figure 4.**
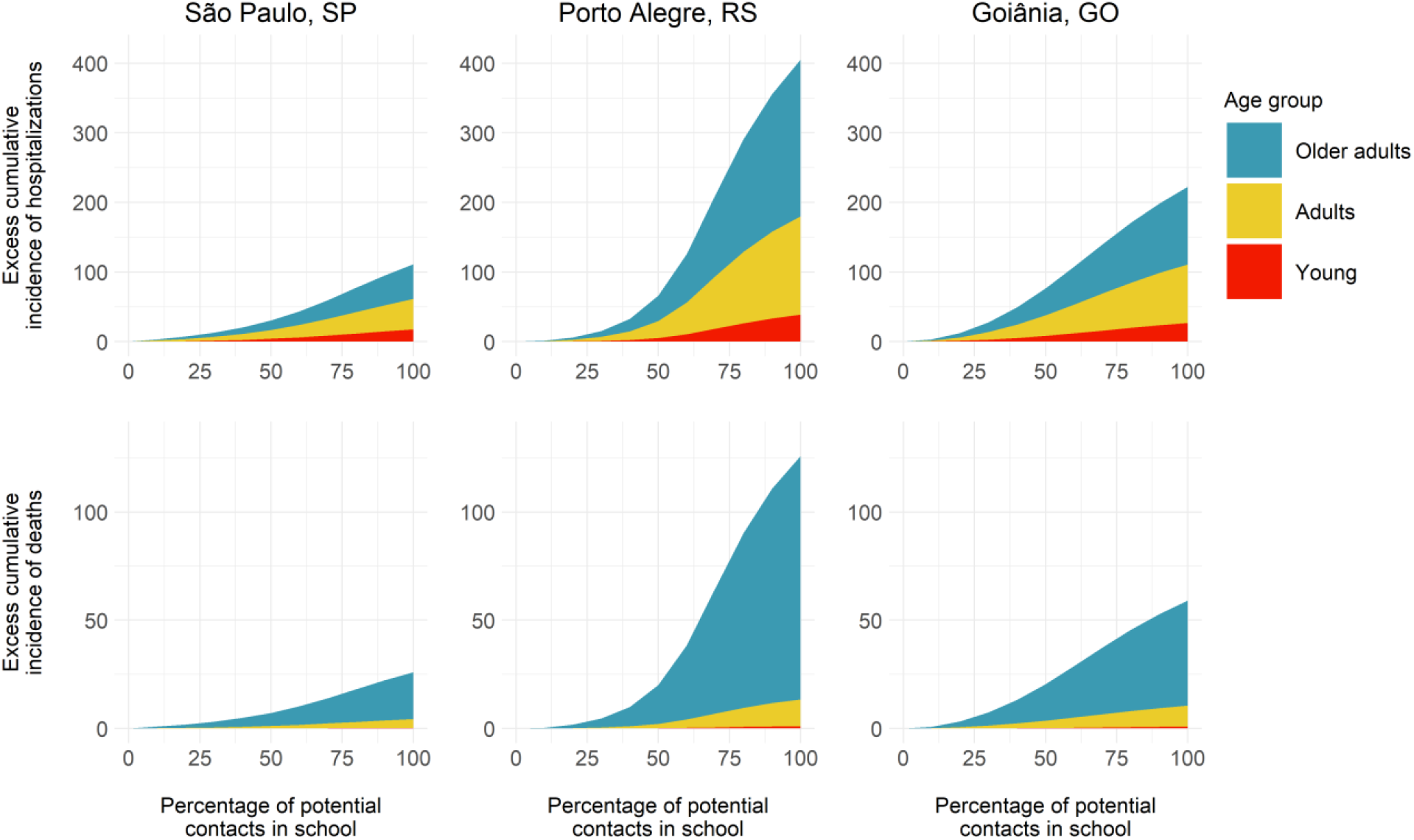
Excess cumulative incidence of COVID-19 hospitalizations and deaths according to the percentage of potential contacts in schools after school reopening.

São Paulo had the smallest excess incidence in cases and deaths with increasing PCS, reaching approximately 100 cases per 100k inhabitants and 25 deaths per 100k inhabitants. On the other hand, Porto Alegre had a considerable increase in incidence, reaching 400 cases per 100 inhabitants and 120 deaths per 100k inhabitants. With PCS values above 50%, Porto Alegre shows a substantial increase in the incidence of cases and deaths. Goiânia presented an intermediate value among the other capitals, reaching just over 200 cases per 100k inhabitants with the total reopening of schools, and a little over 50 deaths per 100k inhabitants. In general, while PCS equal to or below 30% had only a marginal effect on the outcomes, levels of school reopening above this limit resulted in greater increases in the final number of cases and deaths. Despite most contacts on the transmission in school occurring between individuals of young age, by the end of the simulations, they represent less than 12% of cases in all capitals and less than 1% of deaths. On the other hand, older adults comprise a substantial portion of the population infected due to school reopening, representing nearly half of the cases and more than 80% of deaths by the end of the period.

### 3.2 Modelling case isolation, contact tracing, and quarantining scenarios

Finally, our simulations indicate that the implementation of case isolation, contact tracing, and quarantining contacts can reduce the incidence of COVID-19 cases and deaths throughout the pandemic, and particularly upon school reopening. Nonetheless, a significant number of daily tests are required to significantly impact the incidence of COVID-19 cases and deaths (Figure 5). After a certain threshold, increased testing capacity is no longer able to reduce the incidence of cases or deaths. In São Paulo and Goiânia, a daily number of tests corresponding to 3% of the population is required to result in the maximum reduction of cases and deaths due to the isolation of infected individuals. However, while in São Paulo the decrease in the incidence of cases and deaths is independent of PCS, in Goiânia the effectiveness of this measure is greater for lower levels of school reopening (i.e. lower PSC values) but decreases when PCS reaches its maximum value. In Porto Alegre, a testing rate representing 0.75% of its population is required to achieve the maximum reduction in the number of cases if PCS is below 75%, and up to 2% above that value.

**Figure 5.**
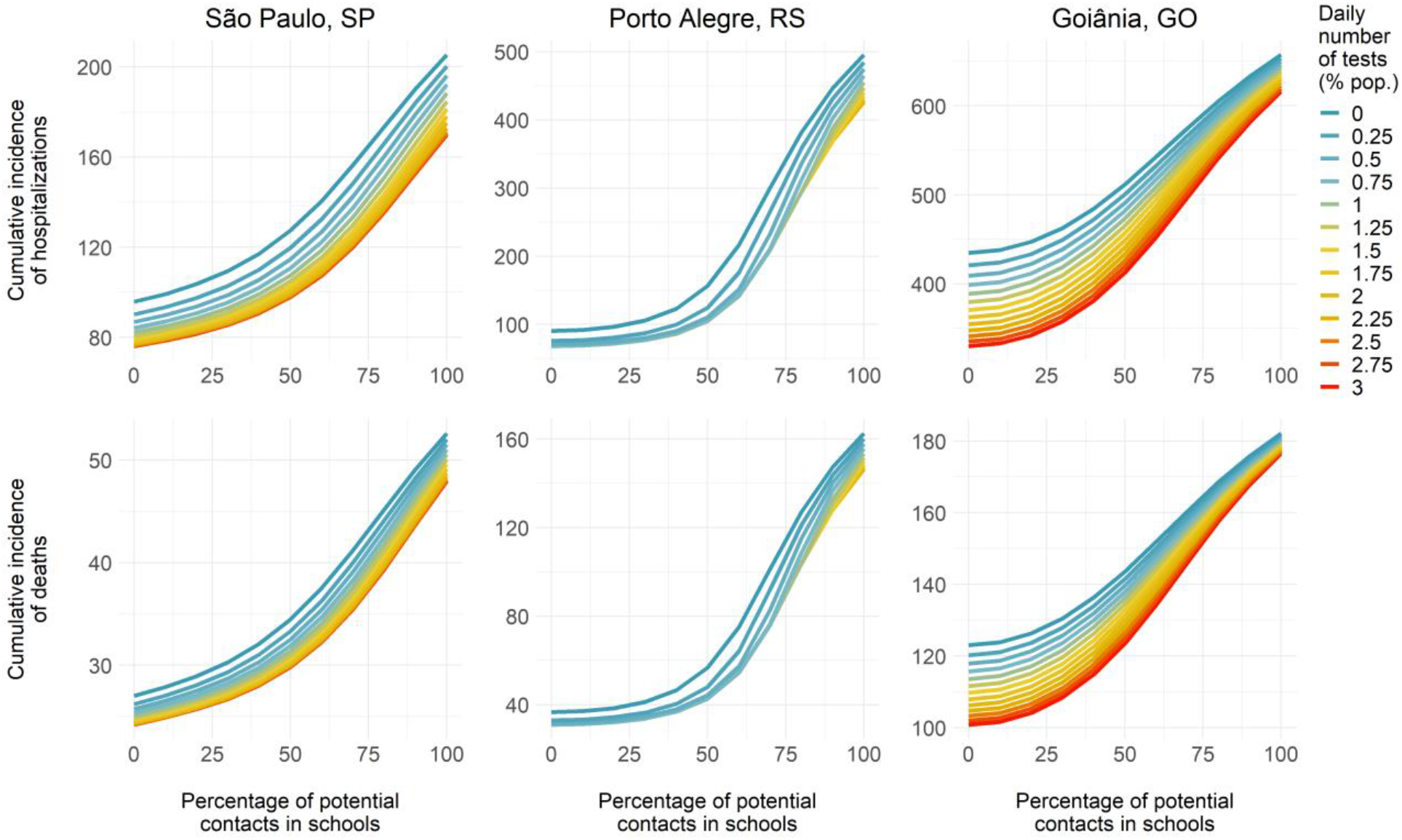
Effects of implementing case isolation, contact tracing, and quarantining contacts in schools on the cumulative incidence of COVID-19 cases and deaths, by the daily number of tests available and scenarios of school reopening (percentage of potential contacts in schools). The projected cumulative incidence of cases and deaths presented is for the period from school reopening (February 1st, 2021) to the end of 2021 (December 31, 2021).

### 3.3 Sensitivity Analysis

For the sensitivity analysis, we evaluated how changes in a parameter of interest can qualitatively and quantitatively alter the simulation results for the different scenarios evaluated for the reopening of schools. Thus, we compared the final difference in the incidence of cases and deaths to a baseline scenario without school reopening. The simulations were repeated for the different school reopening values (PCS) evaluated in the school reopening scenario and compared with the original simulation (supplementary material, section 5). As shown in Figure 6, there is a small variation between the difference in the incidence of cases and deaths for the main results and the outcomes of the model for the sensitivity analysis for the different parameters. In Goiânia, the incidence of cases and deaths is close to the original projections for all PCS values. In São Paulo and Porto Alegre, increasing PCS results in greater variation in the final incidence projections, but the qualitative aspects remain similar to the main results.

**Figure 6.**
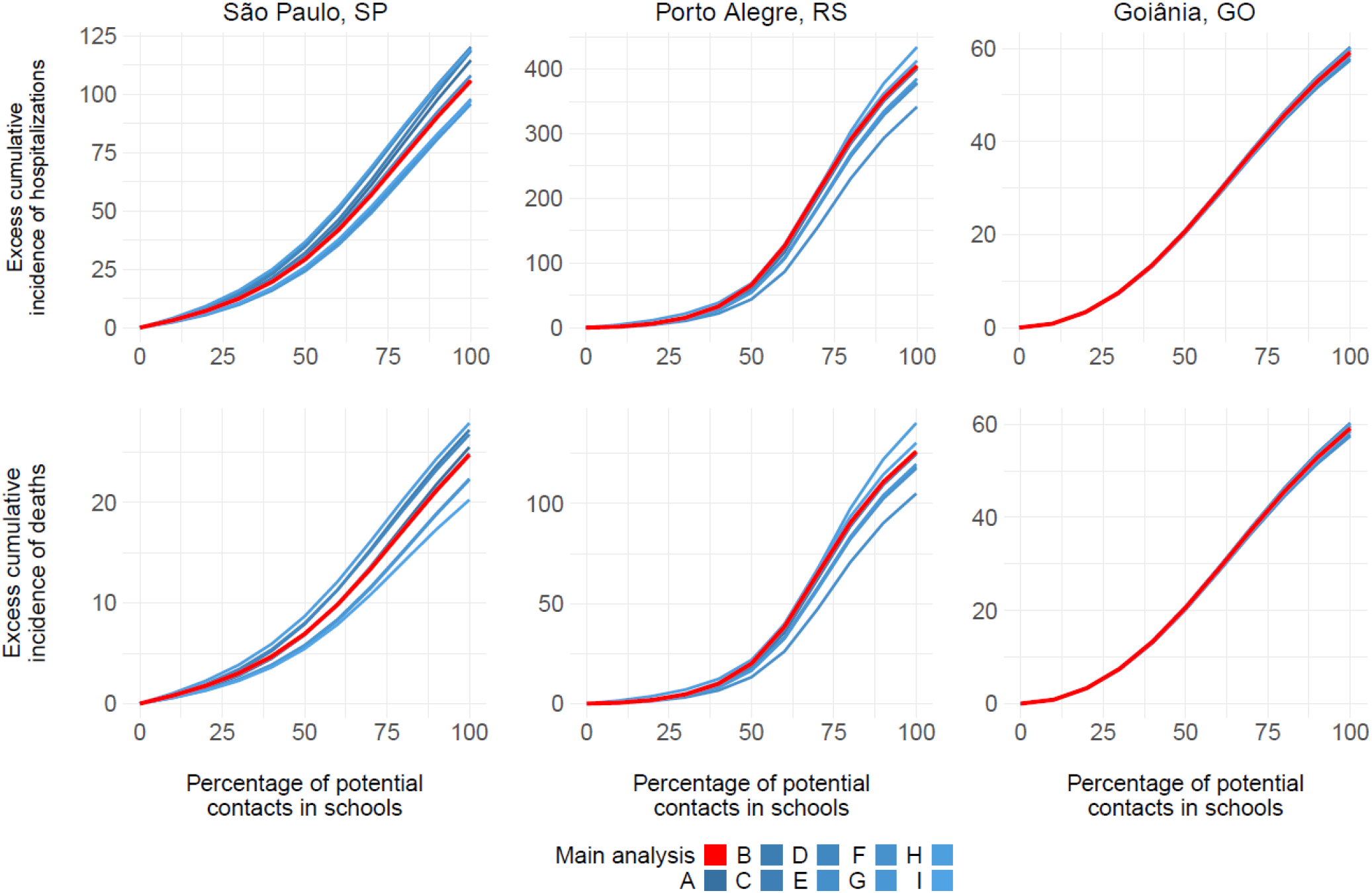
Results of Sensitivity Analysis presenting excess incidence of hospitalizations and deaths according to the percentage of potential contacts in schools after school reopening, when compared to schools fully closed. Different lines represent the models simulated with the parameters fitted on the sensitivity analysis. A; Relative infectiousness of incubation phase (**rho**); B. Relative percentage of regular daily contacts when hospitalized (**rhos**); C. Probability upon infection of developing clinical symptoms for young individuals (**pclin young**); D. Estimated reduction of contact due to mask use (**mask eff**); E. Estimated reduction of contact due to self-isolation if symptomatic (**self eff**); F. Adherence to self-isolation (**selfis cov**); G. Adherence to social distancing at community level (**dist cov**); H. Adherence to home-office (**work cov**); and I. Adherence to cocooning of older adults (**coccoon cov**).

### 3.4 Modelled incidence vs observed incidence

When contrasting the projected COVID-19 incident cases and deaths over time after school reopening, and the true epidemic curves of COVID-19 cases, hospitalization, and deaths in the study sites, the latter strongly surpassed the original projections (Figure 7). As of Abril 2021, in all study sites, the third wave of the pandemic was observed, mostly driven by the predominance of P1 (now denominated *Gamma*) variant of concern, which was first reported in the country in December 2020 in the Northern Region (Manaus), and then disseminated to the whole country. This variant is more transmissible and partially escapes from prior infection and vaccine-induced immunity. Added to this unforeseeable event, several states progressively lifted NPIs early in 2021, considering past trends of disease and confidence in future vaccination.

**Figure 7.**
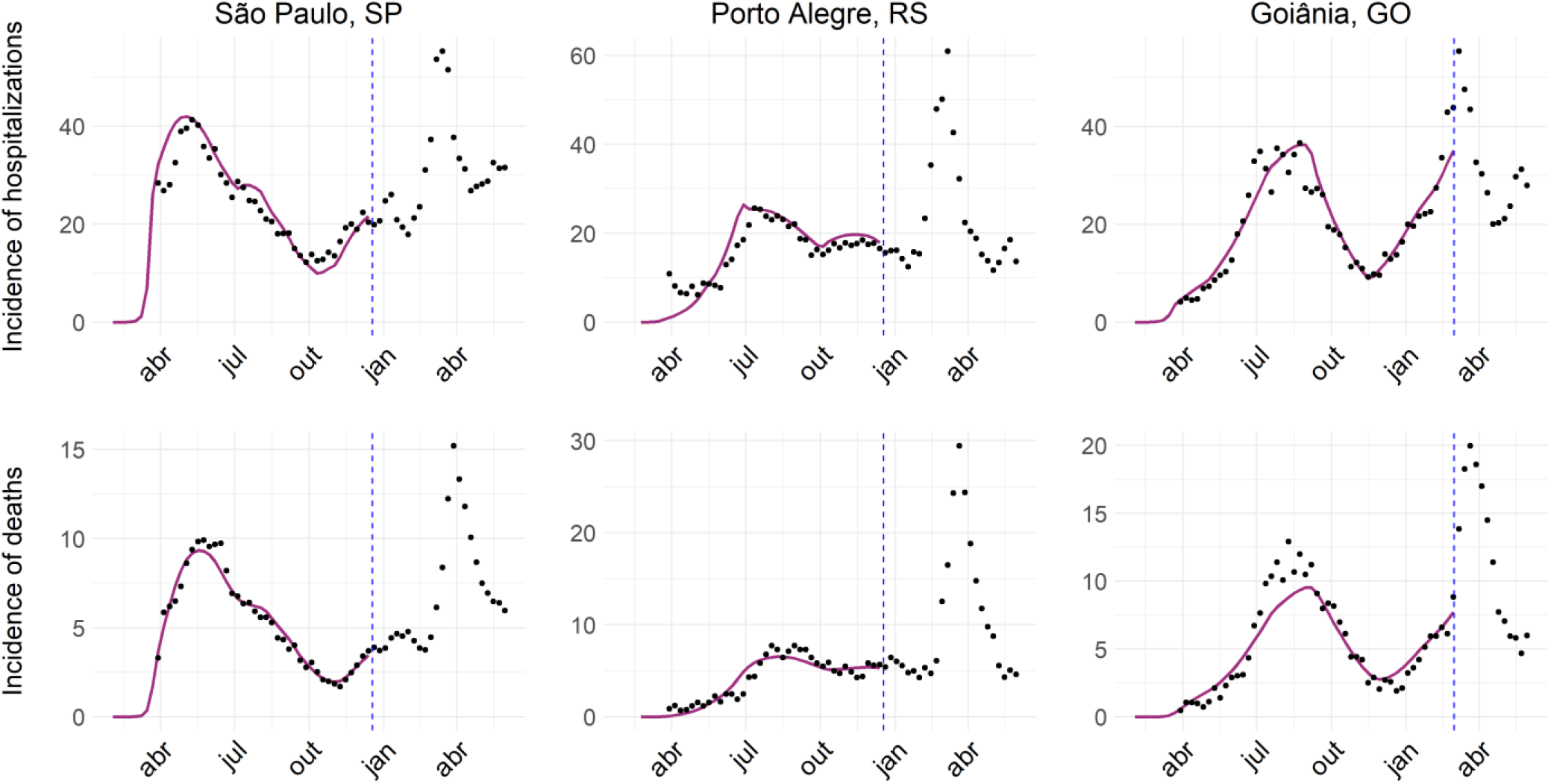
Epidemic curves of hospitalization and deaths for São Paulo, Porto Alegre, and Goiânia. Dots represent the incidence reported for each city. Purple line: simulation from the model. Dashed blue line: maximum date used for fitting.

## 4. Discussion

Among the various recommended non-pharmaceutical interventions to reduce transmission and mitigate COVID-19 pandemic, school closure has been adopted globally, mainly during the first wave of the pandemic in early 2020. In addition to the loss of learning, higher exposure to domestic violence and child abuse, lack of access to meals and immunization delivered at school, social and emotional impacts, among others, are potential impacts of school closures (The Lancet, 2021).

Although most developed countries gradually reopened schools, implementing strict in-school transmission prevention measures, this did not happen in developing and low-income countries. In these countries, schools have been closed for a very long time, with severe short and long-term implications. These are also countries where access to vaccines is limited and vaccination rollout is slower, posing additional challenges to safe school reopening.

School closures were among the first actions taken early in the pandemic in most Brazilian states, even before demonstrated community transmission of SARS-CoV-2 (Silva et al., 2020). Further, Brazil delayed the reopening of schools and stands among the countries in which schools remained closed for the most prolonged period since its inception in 2020.

Considering the continental dimensions and regional specificities of Brazil, it was expected that the epidemiologic pattern of COVID-19 progression would vary significantly among the country’s 27 states, as it did during 2020. This was further challenged by a struggle to define responsibilities at all levels of government (municipal, state, and federal), coupled with erratic and many times misguided communication strategies (Dall’Alba et al., 2021). In the absence of coordinated and equitable responses, the epidemic in Brazil resulted in high and unequal infection and mortality burdens (Castro et al., 2021).

During the first wave of infection in Brazil, COVID-19 cases and deaths were mainly concentrated in large metropolitan areas, but nonetheless, schools in the whole country were closed and remained closed throughout the year 2020. This provided an opportunity to model school reopening in different metropolitan areas with varying epidemiologic, socio-economic, and demographic characteristics, mimicking different LMIC settings facing similar challenges. Our results provide valuable evidence on the potential impact of school reopening during the pandemic and provide insights on when and how to reopen schools more safely in LMIC settings.

First, we demonstrate that the impact of school reopening in terms of incident Covid-19 cases and deaths is small if the potential contacts in school upon reopening are lower than a threshold, that can be as large as 40% (Porto Alegre, RS), becoming more significant when contacts are increased by more than that, e.g., 60% in the previous case. This reinforces the various epidemiologic evidence and recommendations of the need to implement strict prevention and social distancing measures within schools, and not allow for in-person activities for all students at once.

Second, the magnitude of the impact of school reopening in new cases and deaths is dependent on its timing. In settings with declining trends in COVID-19 incidence, small impacts were projected after school reopening (as observed in São Paulo), whereas in settings with increasing trends, the magnitude of the projected impact was significant (as projected in Goiânia).

Third, the excess incidence of COVID-19 cases and deaths is most significant in the older adults, a high-risk group for COVID-19 complications and death. This finding suggests that upon school reopening, vulnerable populations such as older adults must be protected and isolated to minimize the potential impact.

Finally, large-scale testing of suspected individuals and contact tracing strategies are important to minimize the impact of school reopening but require a significant number of tests to do so. Unfortunately, most countries where schools have been closed the longest are also countries that struggle to access diagnostic tests, have limited personnel in place to adequately implement contact tracing strategies, and are thus less likely to have these strategies implemented.

Varying parameters did not impact our results, as demonstrated in sensitivity analysis, pointing to the robustness of our model and parameter estimates.

In summary, we demonstrate that the impact of reopening schools varies in different settings, primarily due to the timing regarding the epidemic curve at the time of reopening but also due to the percentage of potential contacts in school. We considered data adjusted for the period from March through December 2020, and modelled school reopening occurring hypothetically in February 2021. As observed in Goiânia, where the epidemic curve was rising at this time, even with a small percentage of potential contacts at school we observed a new surge. Alternatively, in São Paulo and Porto Alegre, where the curve was declining, it would be possible to reopen schools with a higher percentage of potential contacts (e.g. around 80% and 60% to São Paulo and Porto Alegre, respectively) before observing a trend reversal in the disease incidence. Hence, the epidemiological scenario is important to decide when reopening schools and what percentage of potential contacts are allowed.

It is noteworthy that our modelling was conducted before P1 variant (gamma) circulation in Brazil. Despite the reports of gamma variant circulating in a restricted area of the country at that time (Manaus, in the Amazon Region), it was not possible to model a scenario of school reopening in the context of gamma variant predominance. Since early 2021, the gamma variant emerged and disseminated throughout the country, quickly becoming the predominant circulating variant in all states (Fundação Oswaldo Cruz, 2021). This culminated in synchronic waves of disease in most states of the country in April 2021, and an overload of the already-stretched healthcare system, resulting in a current collapse of the country’s health services and increased mortality from COVID-19 and other causes (Andrade, 2020; Ortega and Orsini, 2020).

School reopening was further delayed as a result and only by the end of May 2021, schools indeed reopened in most states in the country. Nonetheless, when contrasting the projected/modelled impact of school reopening with the additional COVID-19 burden as a result of the third wave of disease-associated to gamma variant emergence and dissemination, in the first quarter of 2021, the estimated impact of school reopening was negligible (Figure 7).

Interestingly, when schools did reopen by the end of May 2021, differently than what had been predicted by several studies (Cruz et al., 2020; España et al., 2021), we did not observe a substantial increase in cases and COVID-19 related mortality. We hypothesize that this might result from the fact that school reopening might represent a marginal impact on new COVID-19 cases and deaths in an epidemic context in which the other measures of social distancing had already been relaxed or lifted (Prem et al., 2020).

Approximately 20 months after the identification of the sustained community transmission of SARS-Cov-2 virus infections on a global scale, there is still some controversy about the risks and benefits of school closure as a public policy to mitigate COVID-19 pandemic. Considering the various short and long-term impacts of long school closures, particularly in LMICs, school reopening should be a societal priority.

Our results provide valuable evidence to policymakers, particularly regarding the safe opening of schools. This can be safely done with social distancing measures in place, at an adequate timing (i.e., when infection transmission in the community is low), progressively (i.e., not bringing all children to in-person activities at once) and associated with strong diagnostic and contact tracing programs in the communities. School reopening must be adapted to the local epidemiologic context and be guided by epidemiologic indicators and should be prioritized among different non-pharmaceutical interventions lifting during the COVID-19 pandemic.

## Supporting information

Model equations; parameterization; fitting procedure; sensitivity analysis

## Data Availability

All data produced are available online at https://github.com/covid19br/school_reopening_manuscript.

