## Supplementary material for "Modeling the impact of school reopening and contact tracing strategies on COVID-19 dynamics in different epidemiologic settings in Brazil": Model equations; parameterization; fitting procedure; sensitivity analysis

[1] *Observatório Covid-19 BR*
[2] *Instituto de Física Teórica, Universidade Estadual Paulista, São Paulo, SP, Brazil*
[3] *Instituto de Patologia Tropical e Saúde Pública, Universidade Federal de Goiás, Goiânia, GO, Brazil*
[4] *Big Data Institute, Li Ka Shing Centre for Health Information and Discovery, Nuffield Department of Medicine, University of Oxford, Oxford, UK*
[5] *Instituto de Ciências Biológicas, Universidade Federal de Goiás, Goiânia, GO, Brazil*
[6] *Instituto de Matemática e Estatística, Universidade Federal do Rio Grande do Sul, Porto Alegre, RS, Brazil*
[7] *Graduate Program in Epidemiology, Universidade Federal do Rio Grande do Sul, Porto Alegre, RS, Brazil*
[8] *Instituto de Biociências, Universidade de São Paulo, São Paulo, Brazil*
[9] *Centro de Matemática, Computação e Cognição - Universidade Federal do ABC, Santo André, Brazil*

#### 1 Introduction

This model framework was first introduced in Aguas et al. [1] and modified to account for brazilian hospital structure and percolation effects in Franco et al. [6]. The code is available at https://github.com/covid19br/school_reopening_manuscript. In Section 2 we introduce our modifications in the Brazilian model structure [6] to account for contact tracing strategies. Section 2.1 describes the equations, along with the explanation and sources of the parameters used. Section 2.2 describes how the force of infection for quarantined and non-quarantined work in the model. Section 2.3 thoroughly describes our contact tracing model. Section 3 lists the interventions used in the main paper. Finally, Section 4 shows the procedure used to fit the model to data and Section 5 describes our approach to sensitivity analysis.

#### 2 Model structure

##### 2.1 Model equations

The model consists in an expanded age-structured SEIR model to account for asymptomatic individuals, a detailed structure of the Brazilian health system, transmission in different settings, and non-pharmaceutical interventions, including contact tracing strategies. We write

$$
\begin{aligned}
\frac{d\mathbf{S}}{dt} &= -\lambda\mathbf{S} + \omega\mathbf{R} + \mathbf{A_G}\cdot\mathbf{S} + \mu_b - \mu_d\mathbf{S} - (Q_{in} + Q_{in,2})\mathbf{S} + Q_d\mathbf{QS} \\
\frac{d\mathbf{E}}{dt} &= \lambda\mathbf{S} - \gamma\mathbf{E} + \mathbf{A_G}\cdot\mathbf{E} - \mu_d\mathbf{E} - (Q_{in} + Q_{in,2})\mathbf{E} + Q_d\mathbf{QE} \\
\frac{d\mathbf{A}}{dt} &= \gamma(1 - P_{clin})(1 - IHR)\mathbf{E} - \nu_i\mathbf{A} + \mathbf{A_G}\cdot\mathbf{A} - \mu_d\mathbf{A} - (Q_{in} + Q_{in,2})\mathbf{A} + Q_d\mathbf{QI} \\
\frac{d\mathbf{I}}{dt} &= (1 - Q_{cov}PT_{cl})\gamma P_{clin}(1 - P_{selfis})(1 - IHR)\mathbf{E} - \nu_i\mathbf{I} + \mathbf{A_G}\cdot\mathbf{I} - \mu_d\mathbf{I} + Q_d\mathbf{QC} \\
\frac{d\mathbf{X}}{dt} &= \gamma P_{selfis}P_{clin}(1 - IHR)\mathbf{E} - \nu_i\mathbf{X} + \mathbf{A_G}\cdot\mathbf{X} - \mu_d\mathbf{X}
\end{aligned}
$$

---

$$
\begin{aligned}
\frac{d\mathbf{H}}{dt} &= \gamma IHR(1 - P_{icu})(1 - H_c)(\mathbf{E} + \mathbf{QE}) - \nu_s\mathbf{H} + \mathbf{A_G}\cdot\mathbf{H} - \mu_d\mathbf{H} \\
\frac{d\mathbf{HC}}{dt} &= \gamma IHR(1 - P_{icu})H_c(\mathbf{E} + \mathbf{QE}) - \nu_{sc}\mathbf{HC} + \mathbf{A_G}\cdot\mathbf{HC} - \mu_d\mathbf{HC} \\
\frac{d\mathbf{ICU}}{dt} &= \gamma IHRP_{icu}(1 - ICU_c)(\mathbf{E} + \mathbf{QE}) - \nu_{icu}\mathbf{ICU} + \mathbf{A_G}\cdot\mathbf{ICU} - \mu_d\mathbf{ICU} \\
\frac{d\mathbf{ICUH}}{dt} &= \gamma IHRP_{icu}ICU_c(1 - ICUH_c)(\mathbf{E} + \mathbf{QE}) - \nu_{icuh}\mathbf{ICUH} + \mathbf{A_G}\cdot\mathbf{ICUH} - \mu_d\mathbf{ICUH} \\
\frac{d\mathbf{ICUC}}{dt} &= \gamma IHRP_{icu}ICU_cICUH_c(\mathbf{E} + \mathbf{QE}) - \nu_{icuc}\mathbf{ICUC} + \mathbf{A_G}\cdot\mathbf{ICUC} - \mu_d\mathbf{ICUC}
\end{aligned}
$$

---

$$
\begin{aligned}
\frac{d\mathbf{R}}{dt} &= \nu_i\mathbf{A} - \omega\mathbf{R} + \nu_i\mathbf{X} + \nu_i\mathbf{I} + \mathbf{A_G}\cdot\mathbf{R} - \mu_d\mathbf{R} + \nu_s(1 - P_dI_{fr})\mathbf{H} - (Q_{in} + Q_{in,2})\mathbf{R} + Q_d\mathbf{QR} \\
&+ \nu_{icu}(1 - P_{dicu}IHFR)\mathbf{ICU} + \nu_{icuc}(1 - P_{dicuc}IHFR)\mathbf{ICUC} + \nu_{sc}(1 - P_{dhc}IHFR)\mathbf{HC} \\
&+ \nu_{icuh}(1 - P_{dicuh}IHFR)\mathbf{ICUH} + \nu_{icuc}(1 - P_{dicuc}IHFR)\mathbf{ICUC}
\end{aligned}
$$

---

$$
\begin{aligned}
\frac{d\mathbf{QS}}{dt} &= -\lambda_q\mathbf{QS} + \omega\mathbf{QR} + \mathbf{A_G}\cdot\mathbf{QS} + \mu_b - \mu_d\mathbf{QS} + (Q_{in} + Q_{in,2})\mathbf{S} - Q_d\mathbf{QS} \\
\frac{d\mathbf{QE}}{dt} &= \lambda_q\mathbf{QS} - \gamma\mathbf{QE} + \mathbf{A_G}\cdot\mathbf{QE} - \mu_d\mathbf{QE} + (Q_{in} + Q_{in,2})\mathbf{E} - Q_d\mathbf{QE} \\
\frac{d\mathbf{QI}}{dt} &= \gamma(1 - P_{clin})(1 - IHR)\mathbf{QE} - \nu_i\mathbf{QI} + \mathbf{A_G}\cdot\mathbf{QI} - \mu_d\mathbf{A} + (Q_{in} + Q_{in,2})\mathbf{A} - Q_d\mathbf{QI} \\
\frac{d\mathbf{QR}}{dt} &= \nu_i(\mathbf{QI} + \mathbf{QC}) + \mathbf{A_G}\cdot\mathbf{QR} - \omega\mathbf{QR} + (Q_{in} + Q_{in,2})\mathbf{R} - Q_d\mathbf{QR} \\
\frac{d\mathbf{QC}}{dt} &= Q_{cov}PT_{cl}\gamma P_{clin}(1 - P_{selfis})(1 - IHR)\mathbf{E} + \gamma P_{clin}(1 - IHR)\mathbf{QE} \\
&- \nu_i\mathbf{I} + \mathbf{A_G}\cdot\mathbf{I} - \mu_d\mathbf{I} - Q_d\mathbf{QC}
\end{aligned}
$$

---

$$
\begin{aligned}
\frac{d\mathbf{C}}{dt} &= r\gamma(1 - IHR)(1 - P_{clin})(\mathbf{E} + \mathbf{QE}) + r_c\gamma(1 - IHR)P_{clin}(\mathbf{E} + \mathbf{QE}) + r_h\gamma IHR(\mathbf{E} + \mathbf{QE}) \\
\frac{d\mathbf{CM}}{dt} &= \nu_sP_{dh}IHFR\mathbf{H} + \nu_{sc}P_{dhc}IHFR\mathbf{HC} + \nu_{icu}P_{dicu}IHFR\mathbf{ICU} + \nu_{icuc}P_{dicuc}IHFR\mathbf{ICUC} \\
&+ \nu_{icuh}P_{dicuh}IHFR\mathbf{ICUH} + \mu_d(\mathbf{H} + \mathbf{HC} + \mathbf{ICU} + \mathbf{ICUC} + \mathbf{ICUH} + \mathbf{I} + \mathbf{X}) \\
\frac{d\mathbf{CMC}}{dt} &= \nu_{sc}P_{dhc}IHFR\mathbf{HC} + \nu_{icuc}P_{dicuc}IHFR\mathbf{ICUC} \\
&+ \nu_{icuh}P_{dicuh}IHFR\mathbf{ICUH} + \mu_d(\mathbf{HC} + \mathbf{ICUC})
\end{aligned}
$$

where each of the dynamic variables (corresponding to the compartments shown in Table 1) is further subdivided in 19 age classes consisting of 5 years age bins (0-4,5-9, up to 90+). Thereby, each of the parameters written in the model, aside from $\mathbf{A_G}$ (ageing matrix), should be thought of as diagonal matrices containing parameter values corresponding to each age class. Take, as an example, the natural mortality rate, given by

$$
\hat{\mu_d} = \mathrm{diag}(\mu_{d1}, \mu_{d2}, ..., \mu_{dD}) = \mathrm{diag}(\vec{\mu_d}).
$$

Note that, in the system of equations presented above, we drop the hats/bolds from all diagonal matrices to avoid an overloaded notation, but keep them in all variables. Thus, each of them actually represents $D = 19$ different ODEs, and therefore the number of equations is $D$ multiplied by the number of compartments. A description of each parameter from the model is available at table 2.

Finally, $\mathbf{A_G}$ implements ageing of the population, and it is defined as a $19 \times 19$ matrix given by:

$$\mathbf{A_G} = \frac{1}{1826.25} \begin{pmatrix} -1 & 0 & 0 & 0 & \dots & 0 & 0 & 0 \\ 1 & -1 & 0 & 0 & \dots & 0 & 0 & 0 \\ 0 & 1 & -1 & 0 & \dots & 0 & 0 & 0 \\ \vdots & & & & \ddots & & \vdots & \\ 0 & 0 & 0 & 0 & & 1 & -1 & 0 \\ 0 & 0 & 0 & 0 & \dots & 0 & 1 & 0 \end{pmatrix} \tag{1}$$

where the denominator accounts for the time to transition between age bins in a 5-year division in units of days.

#### 2.2 Force of infection

Our model assumes two different forces of infection, one for the non-quarantined individuals $\lambda$ and other for quarantined individuals $\lambda_q$. Non-quarantined individuals can be infected by non-quarantined infected individuals in four locations: school, work, home, and in the community. They can also be infected by interacting with quarantined familiars in the "home" setting, and quarantined individuals in other households through the "community" matrix setting. The later considers that transmission by occasional contacts with quarantined individuals in their households may occur, such as in food delivery contexts, for instance. Assuming that $\hat{c}$ is the total contact matrix with percolation effect and non-pharmaceutical interventions described in Franco et al. [6], and $\hat{c}_i$ being the other matrices with the "cocooning of older adults" intervention, with $i = \{\text{home, school, work, community}\}$, we have:

$$\begin{aligned} \lambda = {} & (1 - mask(t))p\hat{c}(\rho\mathbf{E} + \mathbf{A} + \mathbf{I} + imports + \rho_s(\mathbf{H} + \mathbf{ICU} + \mathbf{ICUH}))/\mathbf{P} \\ & + (1 - mask(t))p(1 - f_{perc})\hat{c}_{home}(\rho\mathbf{QE} + \mathbf{QI} + \mathbf{QC} + \mathbf{X} + \mathbf{HC} + \mathbf{ICUC})/\mathbf{P} \\ & + (1 - mask(t))p(1 - Q_{eff,com})\hat{c}_{com}(\rho\mathbf{QE} + \mathbf{QI} + \mathbf{QC} + \mathbf{X} + \mathbf{HC} + \mathbf{ICUC})/\mathbf{P} \end{aligned} \tag{2}$$

where $Q_{eff,com}$ is a parameter of reduction in mean contacts between quarantined and non-quarantined by the "community" contact matrix, *imports* is the value of new imported cases added by day (see Section 3 for details), and $f_{perc}$ and $mask(t)$ are the percolation effect and the usage of mask intervention, respectively, described in Franco et al. [6].

Similarly, a quarantined susceptible individual can be infected by an infected person inside the household, or be infected by interacting through the "community" contact matrix, as follows:

$$\begin{aligned} \lambda_q = {} & (1 - mask(t))p(1 - f_{perc})\hat{c}_{home}(\rho\mathbf{QE} + \mathbf{QI} + \mathbf{QC} + \mathbf{X} + \mathbf{HC} + \mathbf{ICUC})/\mathbf{P} \\ & + (1 - mask(t))p(1 - Q_{eff,com})\hat{c}_{com}(\rho\mathbf{E} + \mathbf{A} + \mathbf{I} + imports + \rho_s(\mathbf{H} + \mathbf{ICU} + \mathbf{ICUH}))/\mathbf{P} \end{aligned} \tag{3}$$

#### 2.3 Contact tracing

To implement the contact tracing strategy, we assume that individuals from compartment $i$, where $i = \{\mathbf{S}, \mathbf{A}, \mathbf{I}, \mathbf{HC}, \mathbf{ICUH}, \mathbf{ICUC}, \mathbf{R}\}$, can be transferred to their respective "quarantined" compartments where their remain isolated, thus, decreasing the chance of infecting other individuals. Isolation occurs after being positively diagnosed as infected by testing, or traced as a secondary contact of a positively diagnosed individual. For simplicity, we refer to all individuals isolated by the contact tracing strategy as "quarantined". Our model supports two ways of testing, one fixing the probabilities $PT_i$ for each compartment or supplying a number of tests applied per day $n_t$. While the implementation of the first case is trivial, for the second one, we first calculate the entrance rate $F_i$ from the exposed (quarantined and non-quarantined) compartment to the compartment studied (for example, $F_H = \gamma IHR(1 - P_{icu})(1 - H_c)(\mathbf{E} + \mathbf{QE})$), with $i$ following the given sequence of priority $i = \{$**ICU, ICUH, H, ICUC, HC, X+CL**$\}$. Then, the probability of testing the compartment $j$ (that follows the same sequence of $i$) is given by:

$$PT_j = \min\left(\max\left(\frac{n_t - \sum_i^j F_i}{F_j + 1}, 0\right), 1\right) \tag{4}$$

where we add 1 to the fraction to avoid division by zero.

Consider again the entrance rate $F_i$, but this time only considering non-quarantined exposed individuals. Then, the entrance rate from a compartment to the quarantined equivalent is given by:

$$Q_{in} = \frac{Q_{cov}\tau_w}{P - Q}\left(\sum_k E_k\hat{c}_k\right)\sum_j PT_j F_j \tag{5}$$

where $Q_{cov}$ is the adherence to quarantine, $\tau_W$ is the time window of traced contacts, $P - Q$ is the total (alive) population discounted for already quarantined individuals, and $E_k$ is the effectiveness of the contact tracing in each $k$ contact matrix (For the results concerning this paper, the only non-zero effectiveness is the one related to school contacts). Notice that the entrance rates are age stratified, thus, the entrance rate to quarantine is also stratified.

Finally, if there are still tests available, they are applied to asymptomatic, exposed, recovered, and susceptible individuals who were identified as secondary contacts of already tested individuals:

$$PT_I = PT_E = \min\left(\max\left(O_d n_{t,2}\frac{\mathbf{S}+\mathbf{E}+\mathbf{A}+\mathbf{R}}{Q_{in}+1}, 0\right), 1\right) \quad (6)$$

where $O_d$ is the overdispersion parameter (we assume equal to 1) and $n_{t,2}$ is the number of remaining tests. Then the second order contacts detected are given by:

$$Q_{in,2} = \frac{Q_{cov}\tau_w}{P-Q}\left(\sum_k E_k \hat{c}_k\right) Q_{in}(PT_E\mathbf{E} + PT_I\mathbf{A}) \quad (7)$$

Notice that we do not assume false positives. Therefore, $PT_S$ and $PT_R$ are equal to zero. Table 6 shows the contact tracing parameters assumed for this study.

### 3 List of interventions

Here we describe the interventions used as input of the model, reproducing (with permission) Franco et al. [6]. Tables 7, 8 and 9 comprises all interventions used in the fitting of the model. Figures 1, 2 and 3 show the timeline of these interventions.

- *Self-Isolation*: Symptomatic individuals that do not require hospitalization voluntarily isolate themselves during the time of infection and reduce the chance of infecting others. The beginning and end period of this intervention is defined by $\theta_{selfis}(t)$ and represents the days $t$ when the population adheres to this behavior. The impact of this NPI depends on its adherence to self-isolation $selfis_{cov}$ and estimated reduction in contacts by self-isolation $selfis_{eff}$ values, where

$$P_{selfis} = selfis_{cov}(t) selfis_{eff} \theta_{selfis}(t) \quad (8)$$

- *Social Distancing*: the population avoids or reduces contacts in the community setting ($\hat{c}_{com}$). This intervention comprises reduction of contacts on churches, markets, social events and gatherings, shopping activities, gyms, and others. The beginning and end period of this intervention is defined by $\theta_{dist}(t)$. The impact of this NPI depends on its adherence to social distancing at community level ($dist_{cov}$) and reduction of contacts in the community among those adhering to social distancing ($dist_{eff}$) values, where:

$$dist(t) = dist_{cov}(t) dist_{eff} \theta_{dist}(t); \quad (9)$$

- *Use of masks*: This intervention comprises individual protection measures, given by the adoption of mask usage. The beginning and end period of this intervention is defined by $\theta_{mask}(t)$. The impact of this NPI depends on its adherence to mask usage ($mask_{cov}$) and effectiveness ($mask_{eff}$), where

$$mask(t) = mask_{cov}(t) mask_{eff} \theta_{mask}(t); \quad (10)$$

- *Work from home*: This intervention reduces contacts in the work environment ($\hat{c}_{work}$) as workers perform their activities from their home. The beginning and end period of this intervention is defined by $\theta_{work}(t)$. The impact of this NPI depends on the adherence to home-office ($work_{cov}$) and reduction of contacts at work among those adhering to home-office ($work_{eff}$), where:

$$work(t) = work_{cov}(t) work_{eff} \theta_{work}(t); \quad (11)$$

- *School closure*: This intervention reduces the contacts in the school setting ($\hat{c}_{school}$) due to limitation of in-school activities or school closures. The beginning and end period of this intervention is defined by $\theta_{school}(t)$. The effectiveness of this NPI depends on the adherence to online (not in-person) school activities ($school_{cov}$) and the reduction of contacts in school upon school closure ($school_{eff}$), where:

$$school(t) = school_{cov}(t) school_{eff} \theta_{school}(t); \quad (12)$$

  Note that in the main text, $school_{cov}$ is also referred as $PCS$ (potential contacts in school).

- *cocooning of older adults*: This intervention reduces the contacts to a proportion of the older adult population, given a minimum age $D^{\dagger}$. The beginning and end period of this intervention is defined by $\theta_{cocoon}(t)$.The effectiveness of this NPI depends on the adherence to cocooning of older adults ($cocoon_{cov}$) and reduction of contacts with older adults in all settings as a results of cocooning older adults ($cocoon_{eff}$). Additional details of this implementation is described in Franco et al. [6].

- *Travel ban*: This intervention models the interruption of travel flow from outside the city and the isolation of cases coming from outside, which reduces or eliminate import cases. This intervention is given by:

$$imports = (1 - travel_{eff})mean\_imports \tag{13}$$

  where ($mean\_imports$) is the mean value of imported cases, $travel_{eff}$ the effectiveness of this intervention, and $imports$ the number of new cases that are added to the population per day.

### 4 Model Fitting

To fit the model onto epidemiological data, we used consolidated time series from Severe Acute Respiratory Infection (SARI) hospitalisations and deaths in São Paulo, Goiânia and Porto Alegre from the SIVEP-Gripe database [4] between the dates described in table 10.

In Brazil, SARI case notification is compulsory (leading to high reporting rates) and SARS-CoV-2 is included as a SARI category. Due to the lack of extensive testing, we assume that using only SARS-CoV-2 confirmed cases would lead to an underestimation of the actual number of cases. Hence, we assume that SARI cases are a better approximation to the number of SARS-CoV-2, rather than only cases confirmed by PCR tests. Since SIVEP-Gripe reports only severe cases that require hospitalisation, we fit SARI cases to the sum over all hospitalised compartments of the model.

Following Franco et al. [6], we chose to use weekly time series for new cases and new deaths to avoid carrying past information into future values, which occurs when using time series of cumulative data.

Based on data from SIVEP [4], we were able to estimate the COVID-19 In-Hospital Fatality Rate (IHFR) and Intensive Care mortality rate (ICMR) for each city (Table 3). Other local parameters are described in Table 4, and local demographic rates per age group in Table 5.

To perform a nonlinear least squares fitting of the free parameters ($p$, $T_{perc}$, $h_{steep}$, $startdate$) to the data, we used the Levenberg-Marquardt algorithm implemented in the minpack.lm R package [5].

To fit both new cases ($C$) and new deaths ($D$), we had to account for residuals in different scales. One way to do that was by normalising each of the variables in respect to their total sum. Therefore, the resulting residual ($R$) is given by:

$$R = \frac{\sum(C_{model} - C_{observed})}{\sum C_{observed}} + \frac{\sum(D_{model} - D_{observed})}{\sum D_{observed}}$$

The algorithm minimises the square of this quantity, while evaluating the respective negative log-likelihood and minimising it.

To perform the non-linear optimisation, the algorithm requires a series of initial guesses. We tested a wide range of $startdate$ values (from $2020-01-01$ to $2020-02-24$) and for each one we ran the fitting algorithm using several reasonable initial guesses for the other free parameters. Hence, this method gives us fitted $p$, $T_{perc}$ and $h_{steep}$ for each $startdate$ considered.

With the goal to find a probability distribution for the fitted parameters [2], we selected the run which returned the lowest residual for each $startdate$, with its respective ($p$, $T_{perc}$, $h_{steep}$) set. We then computed the negative log-likelihood for each start date, $L_t$:

$$L_t = N \ln\left(\frac{1}{N}\sum_{i=1}^{N} R^2_{i,t},\right)$$

from which we can derive the probability for each $startdate$, given by

$$P_t = \frac{exp(-L_t + min(\{L_t\}))}{\sum_t exp(-L_t + min(\{L_t\}))}.$$

Finally, maximising the probability (which is equivalent to minimising the negative log-likelihood), we find sets of best fitted parameters for each of the cities considered (See Table 11)

#### 5 Sensitivity analysis

For the sensitivity analysis, we evaluated how changes in a parameter of interest can qualitatively and quantitatively alter the simulation results for the different scenarios evaluated for the reopening of schools. We set each parameter of interest to be fitted together with the main parameters, sampling uniformly the initial conditions in the range described in 12 and choosing the best fit as result (see tables 13, 14 and 15). Each parameter was fit independently of the others. Since the adherence to the NPI varies in time, the parameter with "cov" were varied by a scaling factor, maintaining the variation in time.

We then compared the final difference in the incidence of cases and deaths in relation to a baseline scenario without school reopening. The simulations were repeated for the different school reopening values (PCS) and compared with the original simulation (see main text).

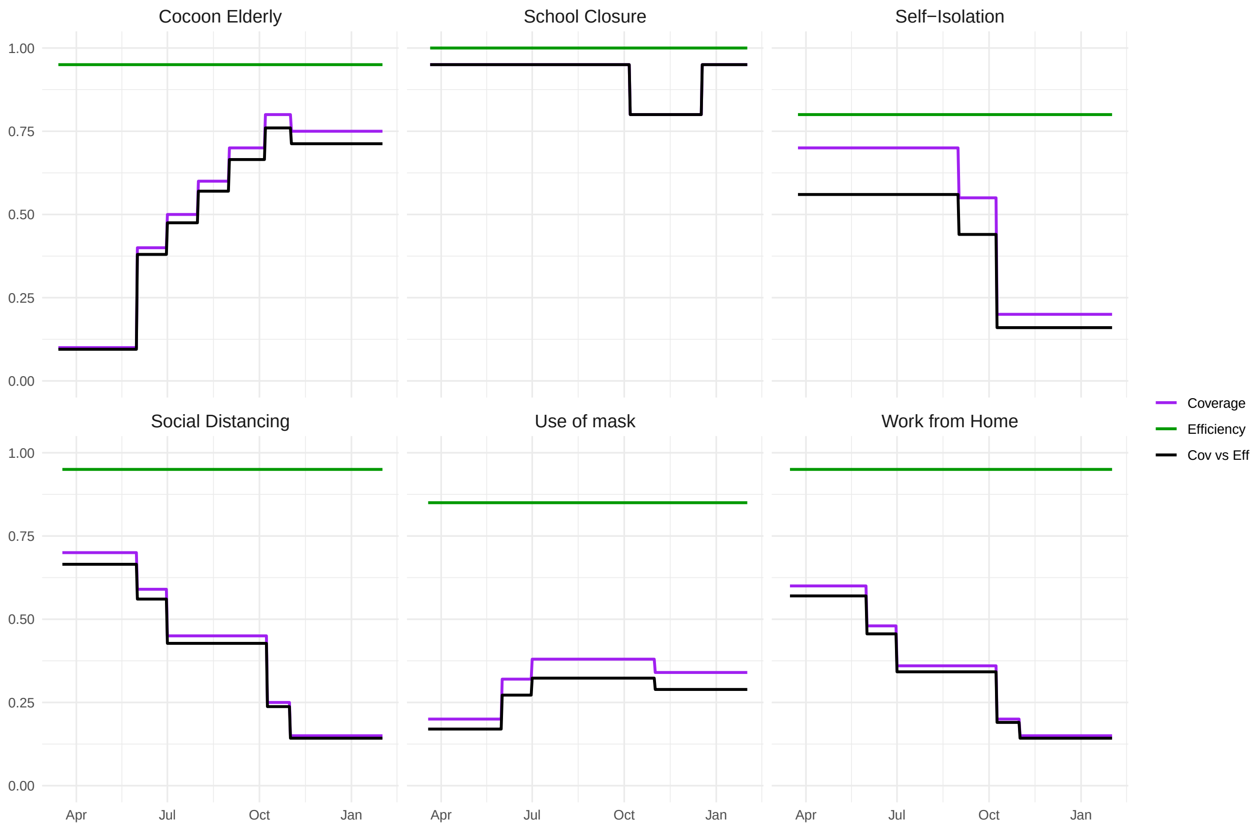


**Figure 1:** Diagram of adherence, reduction of contacts and their product for each of the considered non-pharmaceutical interventions considered in the model for São Paulo, SP.

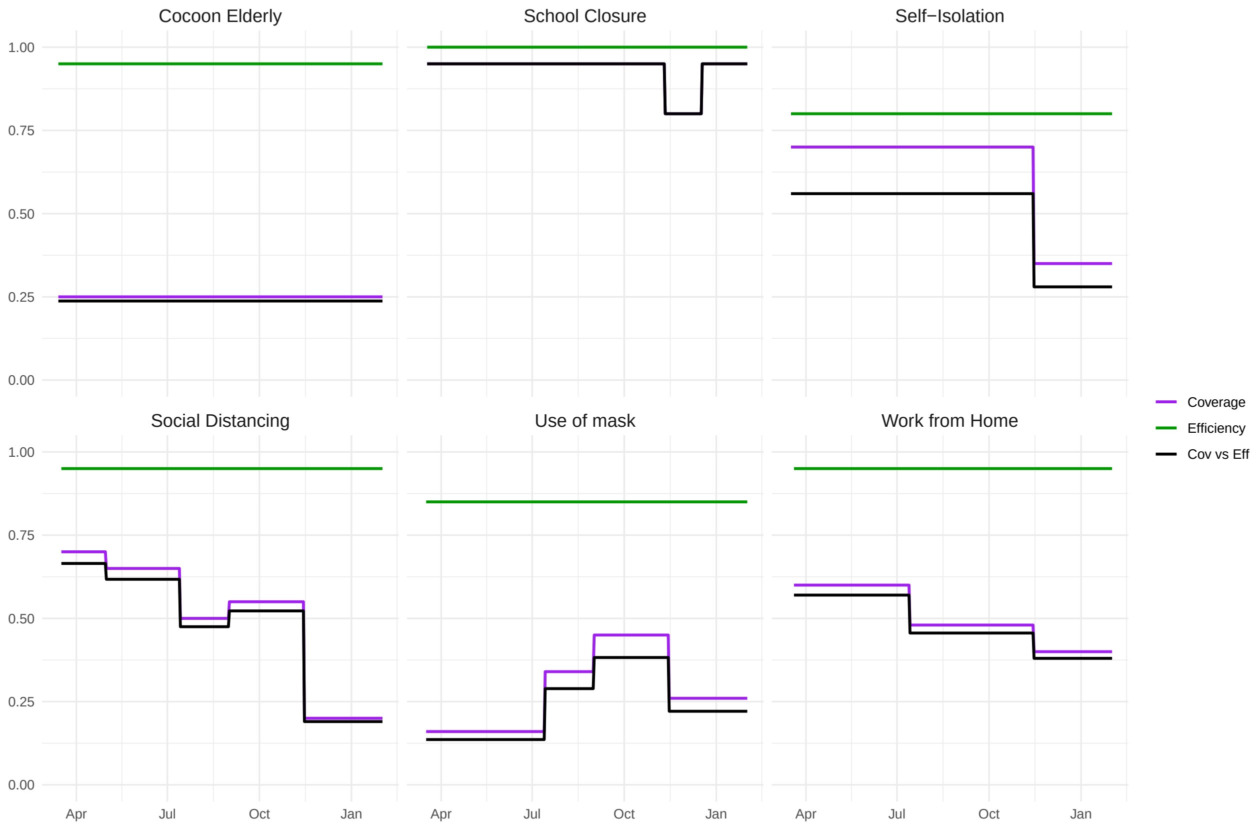


**Figure 2:** Diagram of adherence, reduction of contacts and their product for each of the considered non-pharmaceutical interventions considered in the model for Goiânia, GO.

| code and equations | description |
|---|---|
| **S** | Susceptible population |
| **E** | Infected and presymptomatic population |
| **A** | Infected population, asymptomatic and not isolated |
| **I** | Infected population, mildly symptomatic and not isolated |
| **X** | Infected population, mildly symptomatic and self-isolated at home |
| **H** | Infected population, hospitalized in simple bed. |
| **HC** | Infected population that require hospital treatment but but are denied, due to healthcare system overload |
| **ICU** | Infected population, hospitalised in Intensive Care Units (ICU). |
| **ICUH** | Infected population that require ICU but are hospitalised in simple beds, due to unavailability in ICU beds. |
| **ICUC** | Infected population that require ICU but are denied both an ICU or hospital simple bed, due to healthcare system overload. |
| **R** | Recovered population |
| **QS** | Susceptible population in quarantine |
| **QE** | Infected population in incubation period in quarantine |
| **QI** | Infected asymptomatic population in quarantine |
| **QR** | Recovered population in quarantine |
| **QC** | Mildly symptomatic population in quarantine |
| **C** | Cumulative reported cases |
| $\mathbf{C_M}$ | Cumulative death cases |
| $\mathbf{C_{MC}}$ | Cumulative death cases of critical patients, i.e., those who hospitalization was denied. |

**Table 1:** List of model variables in equations on supplementary material and in the code. Variables written in the main text may be different for readability, here, we stick to the nomenclature used throughout the code to help reproducibility.

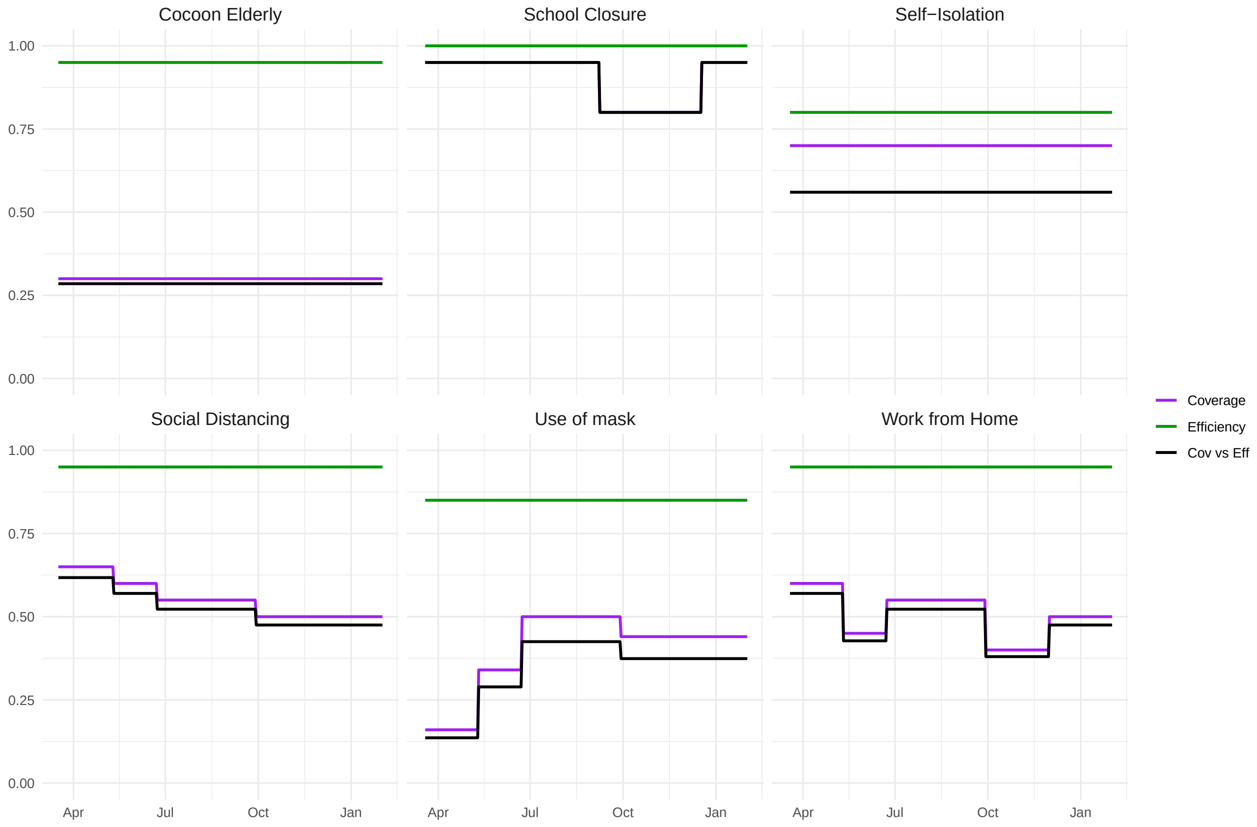


**Figure 3:** Diagram of adherence, reduction of contacts and their product for each of the considered non-pharmaceutical interventions considered in the model for Porto Alegre, RS.

| Code | Equation | Description | Value | Source |
|---|---|---|---|---|
| lam | $\lambda$ | force of infection | Variable | Eq. (2) |
| mort | $\mu_d$ | natural mortality ($days^{-1}$) | See table 5 | IBGE [9] |
| ageing | $\mathbf{A_G}$ | speed of population ageing ($days^{-1}$) | - | Eq. (1) |
| birth | $\mu_b$ | birth rate ($days^{-1}$) | See table 5 | IBGE [7] |
| gamma | $\gamma$ | Inverse average of incubation period ($days^{-1}$) | 1/5.8 | Wei et al. [14] |
| ihr | IHR | Infection hospitalisation rate | See Table 3 | Salje et al. [11] |
| omega | $\omega$ | Rate of which recovered people become susceptible again ($days^{-1}$) | 0 | Assumed |
| rho | $\rho$ | Relative infectiousness of incubation phase | 0.105 | Wei et al. [15] |
| rhos | $\rho_s$ | Relative percentage of regular daily contacts when hospitalized | 0.10 | Assumed |
| pclin | $P_{clin}$ | Probability upon infection of developing clinical symptoms by age groups | 0.305 (0-19)<br>0.560 (20-59)<br>0.690 (60+) | SMSSP [12]<br>Sun et al. [13]<br>Sun et al. [13] |
| selfis | $P_{selfis}$ | Proportion of symptomatic individuals who self-isolate | Variable | Franco et al. [6] (SM) |
| prob_icu | $P_{icu}$ | Proportion of hospitalised individuals who need ICU beds | See Table 3 | Datasus [4] |
| critH | $H_c$ | Proportion of hospitalised individuals who have not received attendance | Variable | Franco et al. [6] (SM) |
| critICU | $ICU_c$ | Proportion of hospitalised individuals who need ICU beds and have not received one | Variable | Franco et al. [6] (SM) |
| critICUH | $ICU_h$ | Proportion of hospitalised individuals who need ICU beds and have not received one and also not have received simple beds | Variable | Franco et al. [6] (SM) |
| nui | $\nu_i$ | Recovery rate of mild symptomatic/asymptomatic individuals ($days^{-1}$) | 1/9 | Cevik et al. [3] |
| nus | $\nu_s$ | Recovery/death rate of hospitalised individuals ($days^{-1}$) | 1/8.3 | Datasus [4] |
| nusc | $\nu_{sc}$ | Recovery/death rate of hospitalised individuals who have not received attendance ($days^{-1}$) | 1/11 | Assumed |
| nu_icu | $\nu_{icu}$ | Recovery/death rate of hospitalised individuals in ICU beds ($days^{-1}$) | 1/14.7 | Datasus [4] |
| nu_icuh | $\nu_{icuh}$ | Recovery/death rate of hospitalised individuals who need ICU beds but received simple beds ($days^{-1}$) | 1/11 | Assumed |
| nu_icuc | $\nu_{icuc}$ | Recovery/death rate of hospitalised individuals who need ICU beds and have not received attendance ($days^{-1}$) | 1/11 | Assumed |
| ifr | IHFR | In hospital fatality rate | See Table 3 | Portella et al. [10] |
| pdeath_h | $P_d$ | Maximum probability of death for a hospitalised infection requiring common bed | See Table 4 | Datasus [4] |
| pdeath_icu | $P_{dicu}$ | Maximum probability of death for a hospitalised infection requiring ICU | See Table 4 | Datasus [4] |
| pdeath_hc | $P_{dhc}$ | Maximum probability of death for a hospitalised infection requiring common bed but not receiving attendance | See Table 4 | Assumed |
| pdeath_icuh | $P_{dicuh}$ | Maximum probability of death for a hospitalised infection requiring ICU but receiving common bed attendance | See Table 4 | Assumed |
| pdeath_icuc | $P_{dicuc}$ | Maximum probability of death for a hospitalised infection requiring ICU but not receiving attendance | See Table 4 | Assumed |
| report | r | Report rate of asymptomatic cases | 0.00 | Assumed |
| reportc | $r_c$ | Report rate of symptomatic cases | 0.01 | Assumed |
| reporth | $r_h$ | Report rate of hospitalized cases | 0.95 | Assumed |

**Table 2:** List of model parameters in equations on supplementary material and in the code. These variables are restricted to epidemiological variables (not the NPI-related ones).

| Age group | Goiânia-GO | | Porto Alegre-RS | | São Paulo-SP | | prob_icu | IHR[1] |
|---|---|---|---|---|---|---|---|---|
| | ICMR | IHFR | ICMR | IHFR | ICMR | IHFR | | |
| 0-4 | 0.29 | 0.034 | 0.26 | 0.028 | 0.14 | 0.014 | 0.45 | 0.1 |
| 5-9 | 0.29 | 0.034 | 0.26 | 0.028 | 0.14 | 0.014 | 0.45 | 0.1 |
| 10-14 | 0.29 | 0.034 | 0.26 | 0.028 | 0.14 | 0.014 | 0.52 | 0.1 |
| 15-19 | 0.29 | 0.034 | 0.26 | 0.028 | 0.14 | 0.014 | 0.52 | 0.1 |
| 20-24 | 0.29 | 0.034 | 0.26 | 0.028 | 0.14 | 0.014 | 0.25 | 0.5 |
| 25-29 | 0.29 | 0.034 | 0.26 | 0.028 | 0.14 | 0.014 | 0.25 | 0.5 |
| 30-34 | 0.25 | 0.036 | 0.21 | 0.013 | 0.2 | 0.028 | 0.32 | 1.1 |
| 35-39 | 0.25 | 0.036 | 0.21 | 0.013 | 0.2 | 0.028 | 0.32 | 1.1 |
| 40-44 | 0.36 | 0.049 | 0.25 | 0.031 | 0.24 | 0.045 | 0.34 | 1.4 |
| 45-49 | 0.36 | 0.049 | 0.25 | 0.031 | 0.24 | 0.045 | 0.34 | 1.4 |
| 50-54 | 0.42 | 0.075 | 0.35 | 0.05 | 0.36 | 0.087 | 0.40 | 2.9 |
| 55-59 | 0.42 | 0.075 | 0.35 | 0.05 | 0.36 | 0.087 | 0.40 | 2.9 |
| 60-64 | 0.59 | 0.166 | 0.56 | 0.109 | 0.52 | 0.162 | 0.48 | 5.8 |
| 65-69 | 0.59 | 0.166 | 0.56 | 0.109 | 0.52 | 0.162 | 0.48 | 5.8 |
| 70-74 | 0.69 | 0.194 | 0.71 | 0.262 | 0.62 | 0.248 | 0.54 | 9.3 |
| 75-79 | 0.69 | 0.194 | 0.71 | 0.262 | 0.62 | 0.248 | 0.54 | 9.3 |
| 80-84 | 0.76 | 0.295 | 0.82 | 0.498 | 0.69 | 0.459 | 0.47 | 26.2 |
| 85-89 | 0.76 | 0.295 | 0.82 | 0.498 | 0.69 | 0.459 | 0.47 | 26.2 |
| 90 + | 0.76 | 0.295 | 0.82 | 0.498 | 0.69 | 0.459 | 0.47 | 26.2 |

**Table 3:** National COVID-19 infection-hospitalization rate (IHR), and COVID-19 In-Hospital Fatality Rate (IHFR) and Intensive Care mortality rate (ICMR) in the 3 study sites, by age sub-groups. Brazil, 2020
[1] Source: Salje et al. [11]

| Parameter | Description | Parameter by site<br>Goiânia<br>GO | <br>Porto Alegre<br>RS | <br>São Paulo<br>SP | Source |
|---|---|---|---|---|---|
| pdeath_h | Probability of death hospitalized infection requiring common bed | 0.295 | 0.498 | 0.459 | Datasus [4] |
| pdeath_icu | Probability of death hospitalized infection requiring ICU | 0.76 | 0.82 | 0.69 | Datasus [4] |
| pdeath_hc | Probability of death hospitalized infection requiring common bed but not receiving attendance | 0.80 | 0.80 | 0.80 | Assumed |
| pdeath_icuh | Probability of death in hospitalized infection requiring ICU but receiving common bed attendance | 0.97 | 0.97 | 0.97 | Assumed |
| pdeath_icuc | Probability of death hospitalized infection requiring ICU | 0.99 | 0.99 | 0.99 | Assumed |
| nus | Duration of hospitalized infection | 7.6 | 9.5 | 8.3 | Datasus [4] |
| nu_icu | Duration of ICU infection | 13.2 | 21.7 | 14.7 | Datasus [4] |
| beds_available | Number common bed | 191 | 1096 | 3000 | Health's Secretary by site |
| icu_beds available | Number ICU bed | 189 | 866 | 5000 | Health's Secretary by site |
| age_distribution | Population by age groups | | | | IBGE [8] |
| | 0-4 | 84000 | 88650 | 768844 | |
| | 5-9 | 87000 | 84793 | 803328 | |
| | 10-14 | 111000 | 76285 | 682355 | |
| | 15-19 | 108000 | 106686 | 750345 | |
| | 20-24 | 123000 | 92060 | 898803 | |
| | 25-29 | 115000 | 109641 | 881006 | |
| | 30-34 | 122000 | 117420 | 983082 | |
| | 35-39 | 131000 | 121941 | 1027565 | |
| | 40-44 | 117000 | 105631 | 955037 | |
| | 45-49 | 104000 | 89660 | 833183 | |
| | 50-54 | 99000 | 87781 | 754688 | |
| | 55-59 | 94000 | 92668 | 678138 | |
| | 60-64 | 79000 | 83814 | 594097 | |
| | 65-69 | 53000 | 68434 | 468480 | |
| | 70-74 | 35000 | 50621 | 340908 | |
| | 75-79 | 25000 | 34142 | 198407 | |
| | 80-84 | 10334 | 16670 | 131116 | |
| | 85-89 | 10334 | 16670 | 72103 | |
| | 90 + | 10334 | 16670 | 48038 | |

**Table 4:** Model Parameter Values used for analysis of COVID-19 school reopening scenarios in Goiânia, Porto Alegre and São Paulo, 2020

| Age groups | Population[1] | Mortality rate deaths/1000 live births[2] | Age groups | Live births[3] |
|---|---|---|---|---|
| 0-4 years | 14789473 | 1445 | | |
| 5-9 years | 14540682 | 118 | | |
| 10-14 years | 15153816 | 143 | 10-14 years | 8853 |
| 15-19 years | 16392753 | 485 | 15-19 years | 201857 |
| 20-24 years | 17285630 | 702 | 20-24 years | 345734 |
| 25-29 years | 17062512 | 726 | 25-29 years | 335131 |
| 30-34 years | 17295219 | 816 | 30-34 years | 295965 |
| 35-39 years | 16675605 | 989 | 35-39 years | 177131 |
| 40-44 years | 14916472 | 1315 | 40-44 years | 42099 |
| 45-49 years | 13288554 | 1874 | 45-49 years | 2437 |
| 50-54 years | 12302879 | 2648 | 50 + years | 226 |
| 55-59 years | 10769470 | 3667 | | |
| 60-64 years | 8831107 | 5016 | | |
| 65-69 years | 6855834 | 6968 | | |
| 70-74 years | 4964070 | 9617 | | |
| 75-79 years | 3387785 | 12497 | | |
| 80-84 years | 2201850 | 50974 | | |
| 85-89 years | 1171537 | 50974 | | |
| 90 + years | 737781 | 50974 | | |

**Table 5:** Demographic data used to calculate the birth and mortality rate in Brazil, 2020.
[1] Source: IBGE [8]
[2] Source: IBGE [9]
[3] Source: IBGE [7]

| Parameter | Description | Value |
|---|---|---|
| $n_t$ | Number of tests available at time $t$ | Varies |
| $n_{t,2}$ | Number of tests available at time $t$ for second order testing | Varies |
| $PT_i$ | Probability of case of compartment $i$ being detected | Varies |
| $Q_{cov}$ | Level of compliance to the contact tracing strategy | 1 |
| $\tau_w$ | Time window of the contact tracing strategy | 2 days |
| $E_{home}$ | Effectiveness of the strategy in tracing contacts from "home" environment | 1 |
| $E_{school}$ | Effectiveness of the strategy in tracing contacts from "school" environment | 1 |
| $E_{work}$ | Effectiveness of the strategy in tracing contacts from "work" environment | 0 |
| $E_{com}$ | Effectiveness of the strategy in tracing contacts from "community" environment | 0 |
| $O_d$ | Overdispersion of the contact tracing strategy | 1 |
| $Q_{eff,com}$ | Effectiveness of quarantine to reduce contacts between quarantined and external individuals | 0.95 |
| $Q_d$ | Inverse of days of quarantine ($days^{-1}$) | 1/10 |

**Table 6:** List of parameters used in the contact tracing model, most are variable and have values described in the main text.

| Self Isolation | | | |
|---|---|---|---|
| Start date | End date | Adherence | Reduction of Contacts |
| 2020-03-24 | 2020-08-31 | 0.70 | 0.80 |
| 2020-09-01 | 2020-10-08 | 0.55 | 0.80 |
| 2020-10-09 | 2021-03-01 | 0.20 | 0.80 |
| Social Distancing | | | |
| Start date | End date | Adherence | Reduction of Contacts |
| 2020-03-18 | 2020-05-31 | 0.70 | 0.95 |
| 2020-06-01 | 2020-06-30 | 0.59 | 0.95 |
| 2020-07-01 | 2020-10-08 | 0.45 | 0.95 |
| 2020-10-09 | 2020-10-31 | 0.25 | 0.95 |
| 2020-11-01 | 2020-03-01 | 0.15 | 0.95 |
| School Closure | | | |
| Start date | End date | Adherence | Reduction of Contacts |
| 2020-03-21 | 2020-10-06 | 0.95 | 1.00 |
| 2020-10-07 | 2020-12-17 | 0.80 | 1.00 |
| 2020-12-18 | 2021-03-01 | 0.95 | 1.00 |
| Use of Mask | | | |
| Start date | End date | Adherence | Reduction of Contacts |
| 2020-03-19 | 2020-05-31 | 0.20 | 0.85 |
| 2020-06-01 | 2020-06-30 | 0.35 | 0.85 |
| 2020-07-01 | 2020-10-31 | 0.42 | 0.85 |
| 2020-11-01 | 2020-03-01 | 0.37 | 0.85 |
| Work from Home | | | |
| Start date | End date | Adherence | Reduction of Contacts |
| 2020-03-16 | 2020-05-31 | 0.60 | 0.95 |
| 2020-06-01 | 2020-06-30 | 0.48 | 0.95 |
| 2020-07-01 | 2020-10-08 | 0.36 | 0.95 |
| 2020-10-09 | 2020-10-31 | 0.20 | 0.95 |
| 2020-11-01 | 2021-03-01 | 0.15 | 0.95 |
| cocooning of older adults | | | |
| Start date | End date | Adherence | Reduction of Contacts |
| 2020-03-14 | 2020-05-31 | 0.10 | 0.95 |
| 2020-06-01 | 2020-06-30 | 0.40 | 0.95 |
| 2020-07-01 | 2020-07-31 | 0.50 | 0.95 |
| 2020-08-01 | 2020-08-31 | 0.60 | 0.95 |
| 2020-09-01 | 2020-10-06 | 0.70 | 0.95 |
| 2020-10-07 | 2020-11-01 | 0.80 | 0.95 |
| 2020-11-02 | 2021-03-01 | 0.75 | 0.95 |
| Travel Ban | | | |
| Start date | End date | Mean imports | Reduction of Contacts |
| 2020-02-19 | 2020-03-18 | 0.20 | 0.0 |
| 2020-03-19 | 2021-03-01 | 0.20 | 0.70 |

**Table 7:** List of interventions used for model fitting in the case of São Paulo, SP.

| Self Isolation | | | |
|---|---|---|---|
| Start date | End date | Adherence | Reduction of Contacts |
| 2020-03-17 | 2020-11-14 | 0.70 | 0.80 |
| 2020-11-15 | 2020-03-01 | 0.35 | 0.80 |
| Social Distancing | | | |
| Start date | End date | Adherence | Reduction of Contacts |
| 2020-03-17 | 2020-04-30 | 0.70 | 0.95 |
| 2020-05-01 | 2020-07-13 | 0.65 | 0.95 |
| 2020-07-14 | 2020-08-31 | 0.50 | 0.95 |
| 2020-09-01 | 2020-11-14 | 0.55 | 0.95 |
| 2020-11-15 | 2020-03-01 | 0.20 | 0.95 |
| School Closure | | | |
| Start date | End date | Adherence | Reduction of Contacts |
| 2020-03-18 | 2020-11-10 | 0.95 | 1.00 |
| 2020-11-11 | 2020-12-17 | 0.80 | 1.00 |
| 2020-12-18 | 2021-01-31 | 0.95 | 1.00 |
| 2021-02-01 | 2021-03-01 | 0.30 | 1.00 |
| Use of Mask | | | |
| Start date | End date | Adherence | Reduction of Contacts |
| 2020-03-17 | 2020-07-13 | 0.16 | 0.85 |
| 2020-07-14 | 2020-08-31 | 0.38 | 0.85 |
| 2020-09-01 | 2020-11-14 | 0.49 | 0.85 |
| 2020-11-15 | 2021-03-01 | 0.29 | 0.85 |
| Work from Home | | | |
| Start date | End date | Adherence | Reduction of Contacts |
| 2020-03-20 | 2020-07-13 | 0.60 | 0.95 |
| 2020-07-14 | 2020-11-14 | 0.48 | 0.95 |
| 2020-11-15 | 2021-03-01 | 0.40 | 0.95 |
| cocooning of older adults | | | |
| Start date | End date | Adherence | Reduction of Contacts |
| 2020-03-14 | 2021-03-01 | 0.25 | 0.95 |
| Travel Ban | | | |
| Start date | End date | Mean imports | Reduction of Contacts |
| 2020-02-19 | 2020-03-18 | 0.20 | 0.0 |
| 2020-03-19 | 2021-03-01 | 0.20 | 0.70 |

**Table 8:** List of interventions used for model fitting in the case of Goiânia, GO.

| Self Isolation | | | |
|---|---|---|---|
| Start date | End date | Adherence | Reduction of Contacts |
| 2020-03-19 | 2020-12-18 | 0.70 | 0.80 |
| Social Distancing | | | |
| Start date | End date | Adherence | Reduction of Contacts |
| 2020-03-17 | 2020-05-10 | 0.65 | 0.95 |
| 2020-05-11 | 2020-06-22 | 0.60 | 0.95 |
| 2020-06-23 | 2020-09-28 | 0.55 | 0.95 |
| 2020-09-29 | 2020-12-18 | 0.50 | 0.95 |
| School Closure | | | |
| Start date | End date | Adherence | Reduction of Contacts |
| 2020-03-19 | 2020-09-07 | 0.95 | 1.00 |
| 2020-09-08 | 2020-12-17 | 0.80 | 1.00 |
| 2020-12-18 | 2021-12-18 | 0.95 | 1.00 |
| Use of Mask | | | |
| Start date | End date | Adherence | Reduction of Contacts |
| 2020-03-19 | 2020-05-10 | 0.16 | 0.85 |
| 2020-05-11 | 2020-06-22 | 0.38 | 0.85 |
| 2020-06-23 | 2020-09-28 | 0.57 | 0.85 |
| 2020-09-29 | 2020-12-18 | 0.49 | 0.85 |
| Work from Home | | | |
| Start date | End date | Adherence | Reduction of Contacts |
| 2020-03-19 | 2020-05-10 | 0.60 | 0.95 |
| 2020-05-11 | 2020-06-22 | 0.45 | 0.95 |
| 2020-06-23 | 2020-09-28 | 0.55 | 0.95 |
| 2020-09-29 | 2020-11-30 | 0.40 | 0.95 |
| 2020-12-01 | 2020-12-18 | 0.50 | 0.95 |
| cocooning of older adults | | | |
| Start date | End date | Adherence | Reduction of Contacts |
| 2020-03-14 | 2020-12-18 | 0.30 | 0.95 |
| Travel Ban | | | |
| Start date | End date | Mean imports | Reduction of Contacts |
| 2020-02-19 | 2020-03-18 | 0.20 | 0.0 |
| 2020-03-19 | 2021-03-01 | 0.20 | 0.70 |

**Table 9:** List of interventions used for model fitting in the case of Porto Alegre, RS.

| City | Start date | End date |
|---|---|---|
| São Paulo | 2020-03-22 | 2020-12-18 |
| Goiânia | 2020-03-22 | 2021-03-05 |
| Porto Alegre | 2020-03-22 | 2020-12-18 |

**Table 10:** Time interval of new hospitalizations from SIVEP-Gripe that were fitted for each city.

| City | Parameter | Estimate | Std. Error | t value | $Pr(>\|t\|)$ |
|---|---|---|---|---|---|
| São Paulo | $p$ | 0.04184 | 0.00010 | 397.8866 | 7.318e-155 |
| | $T_{perc}$ | 0.55151 | 0.00152 | 362.8879 | 4.578e-151 |
| | $h_{steep}$ | 4.58545 | 0.02065 | 222.0186 | 8.082e-131 |
| | startdate | 2020-01-26 | | | |
| Porto Alegre | $p$ | 0.04565 | 0.00019 | 239.8034 | 1.701e-107 |
| | $T_{perc}$ | 0.48442 | 0.34299 | 1.4123 | 0.16209 |
| | $h_{steep}$ | 0.00243 | 0.00062 | 3.8858 | 0.00022 |
| | startdate | 2020-02-18 | | | |
| Goiânia | $p$ | 0.02890 | 5.5e-05 | 523.3179 | 3.664e-166 |
| | $T_{perc}$ | 0.72814 | 0.00307 | 237.4351 | 1.389e-133 |
| | $h_{steep}$ | 15.0106 | 0.05038 | 297.9735 | 6.097e-143 |
| | startdate | 2020-01-27 | | | |

**Table 11:** Best fit results for each of the cities studied.

| | rho | rhos | pclin young | hand eff | hand cov | selfis eff | selfis cov | dist cov | work cov | cocoon cov |
|---|---|---|---|---|---|---|---|---|---|---|
| Min | 0 | 1 | 0.3 | 0.5 | 0.4 | 0.5 | 0.5 | 0.5 | 0.5 | 0.5 |
| Max | 88 | 100 | 0.7 | 0.99 | 1.25 | 0.99 | 1.25 | 1.25 | 1.25 | 1.25 |

**Table 12:** Range of variation of each parameter for the sensitivity analysis.

| startdate | $p$ | $P_{thresh}$ | $P_{steep}$ | SA parameter | Best fit value | Original value | Residual | Neg loglik |
|---|---|---|---|---|---|---|---|---|
| 2020-01-26 | 0.0417 | 0.55 | 4.49 | - | - | - | 0.00208 | -798.33 |
| 2020-01-26 | 0.0372 | 0.60 | 4.64 | rho | 16.38 | 10.50 | 0.00324 | -1011.15 |
| 2020-01-26 | 0.0416 | 0.58 | 4.7 | rhos | 10.15 | 10.0 | 0.00412 | -987.58 |
| 2020-01-26 | 0.0413 | 0.58 | 4.62 | pclin young | 0.56 | 0.305 | 0.00284 | -1023.88 |
| 2020-01-26 | 0.0414 | 0.58 | 4.63 | mask eff | 0.94 | 0.85 | 0.00313 | -1014.5 |
| 2020-01-26 | 0.0416 | 0.57 | 4.74 | self eff | 0.84 | 0.80 | 0.00295 | -1020.17 |
| 2020-01-26 | 0.041 | 0.60 | 4.63 | selfis cov | 1.17 | 1.0 | 0.00278 | -1026.02 |
| 2020-01-26 | 0.042 | 0.61 | 4.58 | dist cov | 1.10 | 1.0 | 0.00297 | -1019.73 |
| 2020-01-26 | 0.0417 | 0.53 | 4.67 | work cov | 0.93 | 1.0 | 0.00302 | -1017.97 |
| 2020-01-26 | 0.0414 | 0.57 | 4.84 | cocoon cov | 1.11 | 1.0 | 0.00377 | -996.1 |

**Table 13:** Sensitivity analysis of the fitting for São Paulo, SP.

| startdate | $p$ | $P_{thresh}$ | $P_{steep}$ | SA parameter | Best fit value | Original value | Residual | Neg loglik |
|---|---|---|---|---|---|---|---|---|
| 2020-02-18 | 0.0453 | 0.50 | 0 | - | - | - | 0.00277 | -776.77 |
| 2020-02-18 | 0.0455 | 0.50 | 0 | rho | 10.78 | 10.50 | 0.00278 | -776.44 |
| 2020-02-18 | 0.0455 | 0.49 | 0 | rhos | 10.14 | 10.0 | 0.00265 | -779.98 |
| 2020-02-18 | 0.0455 | 0.50 | 0.01 | pclin young | 0.3 | 0.305 | 0.00266 | -779.91 |
| 2020-02-18 | 0.0451 | 0.20 | 0.05 | mask eff | 0.81 | 0.85 | 0.00233 | -789.71 |
| 2020-02-18 | 0.0432 | 0.51 | 0.02 | self eff | 0.53 | 0.80 | 0.00220 | -794.24 |
| 2020-02-18 | 0.0446 | 0.49 | 0.05 | selfis cov | 0.87 | 1.0 | 0.00229 | -791.05 |
| 2020-02-18 | 0.0448 | 0.50 | 0 | dist cov | 0.94 | 1.0 | 0.00257 | -782.36 |
| 2020-02-18 | 0.0409 | 0.49 | 1.31 | work cov | 0.52 | 1.0 | 0.00228 | -791.52 |
| 2020-02-18 | 0.0451 | 0.50 | 0 | cocoon cov | 0.83 | 1.0 | 0.00256 | -782.59 |

**Table 14:** Sensitivity analysis of the fitting for Porto Alegre, RS.

| startdate | $p$ | $P_{thresh}$ | $P_{steep}$ | SA parameter | Best fit value | Original value | Residual | Neg loglik |
|---|---|---|---|---|---|---|---|---|
| 2020-01-27 | 0.0287 | 0.73 | 15.0 | - | - | - | 0.00224 | -1012.19 |
| 2020-01-27 | 0.0299 | 0.70 | 22.92 | rho | 9.21 | 10.50 | 0.00280 | -1025.46 |
| 2020-01-27 | 0.0299 | 0.71 | 15.07 | rhos | 6.56 | 10.0 | 0.00280 | -1025.24 |
| 2020-01-27 | 0.0293 | 0.73 | 15.00 | pclin young | 0.590 | 0.305 | 0.00291 | -1021.66 |
| 2020-01-27 | 0.0294 | 0.71 | 15.11 | mask eff | 0.87 | 0.85 | 0.00309 | -1015.6 |
| 2020-01-27 | 0.0303 | 0.71 | 14.38 | self eff | 0.91 | 0.80 | 0.00288 | -1022.57 |
| 2020-01-27 | 0.0293 | 0.72 | 14.5 | selfis cov | 0.99 | 1.0 | 0.00284 | -1024.16 |
| 2020-01-27 | 0.0294 | 0.74 | 14.85 | dist cov | 1.03 | 1.0 | 0.00281 | -1024.89 |
| 2020-01-27 | 0.0291 | 0.71 | 14.92 | work cov | 0.94 | 1.0 | 0.00282 | -1024.65 |
| 2020-01-27 | 0.0296 | 0.71 | 14.96 | cocoon cov | 0.82 | 1.0 | 0.00255 | -1034.54 |

**Table 15:** Sensitivity analysis of the fitting for Goiânia, GO.
